# Use of clustering to improve estimation of epidemic model parameters under a Bayesian hierarchical framework

**DOI:** 10.1101/2022.12.08.22283266

**Authors:** Punya Alahakoon, James M. McCaw, Peter G. Taylor

## Abstract

We study infectious disease outbreaks that have evolved in isolation without the influence of one another. If stochastic effects are identified within each outbreak, it is necessary to model the dynamics with stochastic epidemic models. However, the accuracy of the estimated model parameters depends on several factors including the statistical inference methodologies that are used. One approach to making inferences from multiple outbreak data is the use of a Bayesian hierarchical model. This statistical framework allows simultaneous inference for multiple outbreaks and the estimation of model parameters at a group level. A hierarchical model will generally provide improved estimates; however, we show that this is not always true when the variability among model parameter values of the outbreaks is high. We further show that subsets of outbreaks with similar parameter values can be identified prior to a hierarchical analysis using common clustering algorithms such as k-means. When hierarchical analyses are carried out for these pre-identified subsets of outbreaks, parameter estimates are improved compared to those estimated under a hierarchical analysis for the complete set of outbreaks. We have applied our estimation framework within a simulation-based experiment using synthetic data generated from stochastic *SIRS* models. The framework is generalizable to other biological data.

## 1 Introduction

Early epidemic modeling was largely based on deterministic models (Kermack & McKendrick, 1927, 1932, 1933; Ross, 1911). However, disease dynamics are often affected by stochastic effects that result in the random nature of epidemiological dynamics (M. Bartlett, 1956), stochastic extinction (Andersson & Britton, 2012; M. Bartlett, 1964), and fluctuations that deviate away from deterministic predictions (M. S. Bartlett, 1960; Keeling & Ross, 2008). Stochastic effects are further relevant when the host population is small or disease prevalence is low (Andersson & Britton, 2012; Britton, 2010; Lloyd-Smith et al., 2005). When such effects are observed or anticipated, stochastic models are required for analyses.

Here we consider outbreaks of an infectious disease occurring in multiple communities which we will call sub-populations. If we observe these outbreaks in settings such as on ships, islands, or in hospitals, then it is reasonable to assume that they will evolve without influencing one another. Similarly, non-interaction of individuals within clinical trials will also generate independently evolving within-host dynamics (Cao et al., 2019). Here, we conceive of a hypothetical setting in which such multiple outbreaks of an infectious disease are observed and each outbreak is modeled by a stochastic epidemic model. Our aim is to accurately estimate the model parameters.

Accurate estimates of epidemic model parameters are crucial to understanding disease-related characteristics (McKinley et al., 2018) and thresholds for disease extinction (Allen & Lahodny Jr, 2012). They are further helpful in identifying and studying effective disease control measures (O’Dea, Pepin, Lopman, & Wilke, 2014). Assuming that the proposed epidemic model structure is appropriate for studying the infectious disease in question, the accuracy of parameter estimates relating to multiple outbreaks can depend on various factors, including the availability of data (Capaldi et al., 2012), the accuracy of algorithms that are used to estimate parameters, and statistical modeling and other inferential methodologies that are used (Held, Hens, D O’Neill, & Wallinga, 2019; McKinley et al., 2018). In this study, we focus on the latter.

Hierarchical models, commonly known as mixed effects models or multi-level models in statistics (McElreath, 2020), are ideal to make inferences on multiple outbreak data. A hierarchical model is a statistical modeling framework that can be used to study inherently nested data (Gelman & Hill, 2006; Goldstein, 2011; Royle & Dorazio, 2008) and/or when it is desired to make inferences at multiple levels (population and sub-population level) (J. Kruschke, 2014). These types of frameworks allow multiple and simultaneous comparisons of data (J. Kruschke, 2014; McElreath, 2020). They can improve parameter estimates compared to when each outbreak is studied independently by incorporating information from higher levels for estimation (Alahakoon, McCaw, & Taylor, 2022a, 2022b; J. Kruschke, 2014). Within a hierarchical framework, we assume that there exists a dependence structure across some of the model parameters of the epidemic models. This dependence structure arises by allowing sub-population specific parameters to be sampled from a common distribution (Gelman et al., 2013).

In this study, we consider three groups of multiple outbreaks of an infectious disease where each outbreak takes place in small sub-populations and their dynamics evolve independently from each other. The transmission pattern of each outbreak will be different because of stochastic effects as well as the fact that different sub-populations have different model parameters. We generate synthetic outbreak data from a stochastic *SIRS* model with parameters generated such that the groups have three distinct transmission levels. We first apply a Bayesian parameter estimation algorithm for each outbreak independently to obtain samples from posterior distributions for all the model parameters. Using the posterior samples of the transmission rate, we then apply the k-means clustering algorithm to identify and recover groups of sub-populations that have similar transmission rates under this non-hierarchical analysis. Next, we apply Bayesian hierarchical analyses to these identified groups. We demonstrate that when groups of outbreaks having similar model parameters are identified prior to the hierarchical analysis, parameter estimates of the sub-populations are improved compared to under a hierarchical analysis on all the outbreaks together. We show that this additional step of clustering is practical in terms of computational burden when applied within a Bayesian hierarchical approach.

## 2 Background

### 2.1 The Markovian SIRS model

We denote the number of susceptibles, infectious, and recovered individuals as *S*(*t*), *I*(*t*), and *R*(*t*) respectively at time *t* in a well-mixed closed sub-population of size *N*. We parameterise an *SIRS* model with *β, γ*, and *μ*, the transmission, recovery, and waning immunity rates respectively. We formulate the *SIRS* stochastic system as a continuous-time Markov chain with bi-variate states (*S*(*t*), *I*(*t*)) with the transition rates presented in Table 1. The model structure is shown in Figure 1.

**Table 1:** Transition rates of an *SIRS* model

| Event | Transition | Rates |
| --- | --- | --- |
| Infection | $(s, i) \rightarrow (s - 1, i + 1)$ | $\beta si / (N - 1)$ |
| Recovery | $(s, i) \rightarrow (s, i - 1)$ | $\gamma i$ |
| Loss of immunity | $(s, i) \rightarrow (s + 1, i)$ | $\mu r, (r = N - s - i)$ |

**Figure 1:**
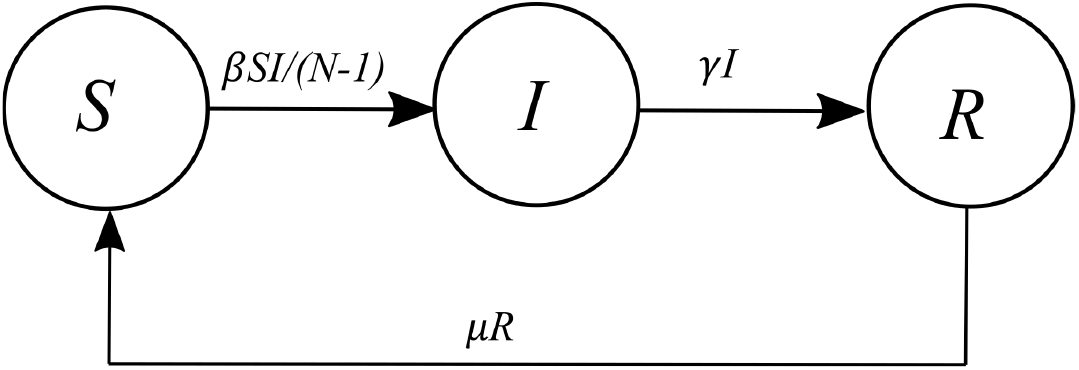
*SIRS* model structure.

### 2.2 A Bayesian hierarchical modeling framework

Let us assume that we are studying a set of outbreaks taking place in *J* sub-populations. We will model each outbreak using a stochastic *SIRS* model as a continuous-time Markov chain with transition rates defined in Table 1. We will denote the model parameter set corresponding to outbreak *j* (*j* = 1, 2, …, *J*) as a vector ***θ***_*j*_ where ***θ***_*j*_ = (*β*_*j*_, *γ*_*j*_, *μ*_*j*_). Under a hierarchical modeling framework, we assume that the model parameters for this set of outbreaks are drawn from a common distribution (Gelman et al., 2013).

Our hierarchical framework is similar to that of Alahakoon et al. (2022a) which has three levels. We use Level I to represent prevalence data, ***y***_*j*_ (for *j* = 1, 2, 3, …, *J*), that are observed at discrete time points. Level II is used to represent the conditional prior distribution *p*(***θ***_*j*_|**Ψ**) that describes the structural relationship between sub-population specific parameters and the hyper-parameters. The hyper-prior distributions, *p*(**Ψ**), are represented in Level III (Alahakoon et al., 2022a).

The joint posterior distribution for a set of outbreaks consisting of *J* sub-populations is,

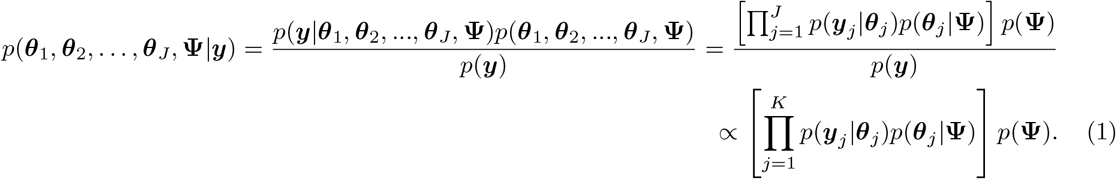

## 3 Materials and Methods

### 3.1 Synthetic data generation

We constructed synthetic data for three groups of sub-populations with each group consisting of 10 sub-populations. The sub-population sizes for sub-groups A, B, and C were 500, 1000, and 1000 respectively. We assumed an observed model in which daily infection prevalence for each sub-population was measured without observational noise over a 30-day period. We assumed that an outbreak in a sub-population started with one infectious individual and generated data accordingly.

One of the objectives of this study is to identify, when present, subsets of outbreaks with similar parameter values. Therefore, we chose the population level (mean) transmission rates of the sub-groups to exhibit this contrast. Accordingly, we chose three transmission levels for the sub-groups: high, low, and medium for Sub-groups A, B, and C respectively.

Transmission, recovery, and waning immunity rates for each sub-group were generated by sampling from truncated normal distributions. Here, TN(*a, b*^2^, *c, d*) is a Truncated Normal distribution with *a* mean, *b* standard deviation, truncated on the interval (*c, d*). We randomly generated *β*_*j*_ (for *j* = 1, 2, …, 10) from TN(3.5, 0.25^2^, 1, 10) for Sub-group A, TN(2, 0.15^2^, 1, 10) for Sub-group B, and TN(2.5, 0.5^2^, 1, 10) for Sub-group C. For all three groups, we used common distributions to generate *γ*_*j*_ and *μ*_*j*_. For *γ*_*j*_, the sampling distribution was TN(1, 0.05^2^, 0, 4) and for *μ*_*j*_, it was TN(0.06, 0.01^2^, 0.04, 4). Table 2 presents the means and standard deviations of the sampled parameters for the groups of sub-populations, and all three groups combined.

**Table 2:** Means and standard deviations of the parameters of the groups

| Group | $\beta_j$ | | $\gamma_j$ | | $\mu_j$ | |
| --- | --- | --- | --- | --- | --- | --- |
|  | Mean | Standard deviation | Mean | Standard deviation | Mean | Standard deviation |
| **Sub-group A** (High transmission) | 3.477 | 0.339 | 1.011 | 0.064 | 0.061 | 0.007 |
| **Sub-group B** (Low transmission) | 1.943 | 0.192 | 1.007 | 0.038 | 0.059 | 0.012 |
| **Sub-group C** (Medium transmission) | 2.674 | 0.414 | 1.013 | 0.054 | 0.056 | 0.008 |
| **One group**<br>(Sub-groups A, B and C) | 2.698 | 0.712 | 1.010 | 0.051 | 0.059 | 0.009 |

After generating parameter sets for all 30 sub-populations, we simulated a single sample path from the stochastic *SIRS* model defined in Section 2 for each sub-population using the Doob-Gillespie (Doob, 1945; Gillespie, 1977) algorithm. When generating a sample path for a sub-population, we repeatedly generated the sample path until an outbreak occurred so that no sub-population exhibited an initial fade-out. We used the same criteria as Alahakoon et al. (2022a) to identify an outbreak (see Supplementary Material S5 for these details). Once a sample path was generated for a sub-population, we recorded the daily prevalence of infections for a 30-day period. Figure 2 illustrates these time series for the sub-populations in each of the three groups.

**Figure 2:**
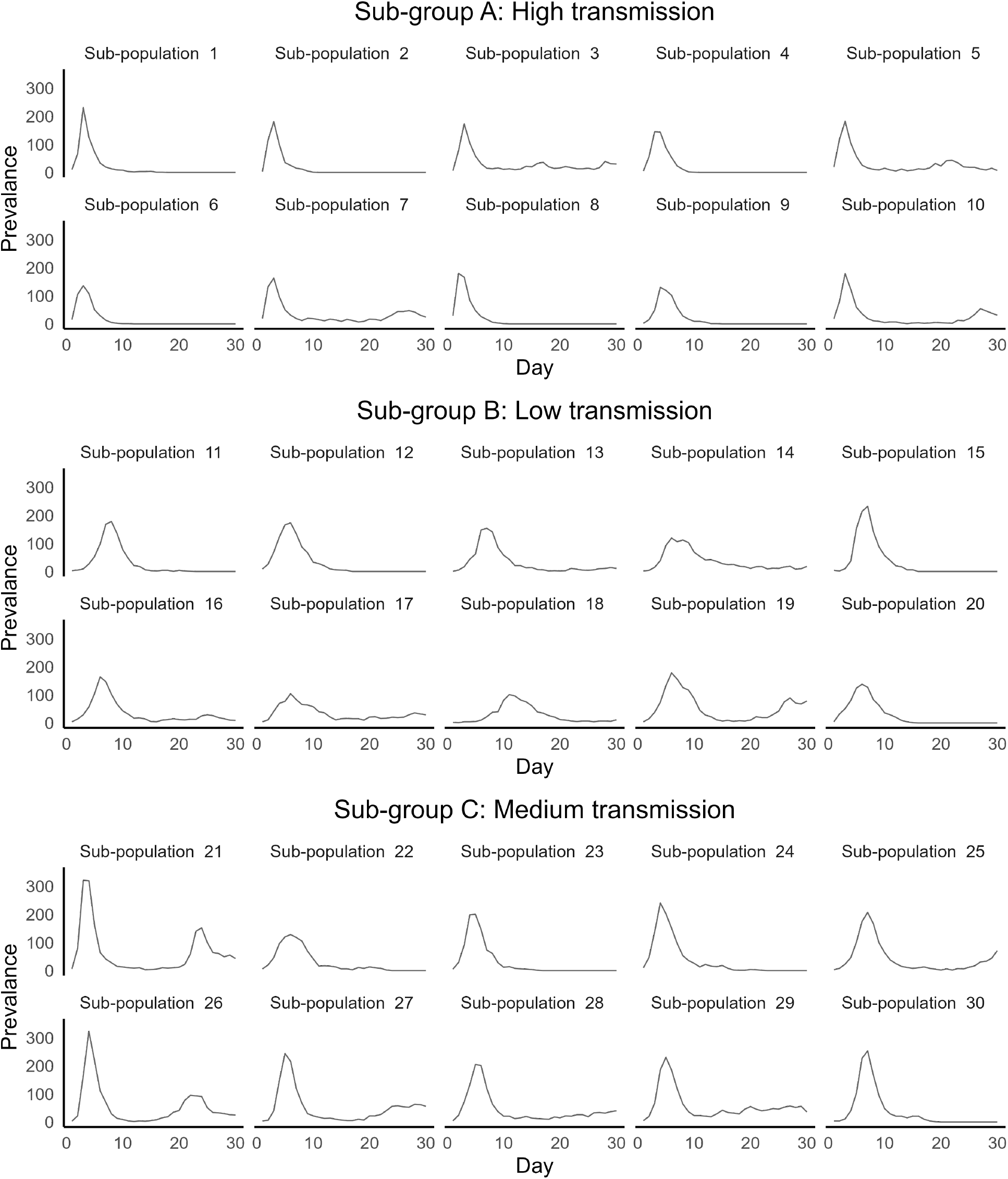
Time-series data for the three sub-groups

### 3.2 Estimation framework

We conducted hierarchical analyses under two alternative scenarios: 1) a hypothetical scenario in which we have perfect knowledge regarding the allocation of sub-populations to each of the three sub-groups. 2) a pragmatic scenario in which we must identify groupings.

Under both scenarios, we conducted the hierarchical analysis using the algorithm introduced by Alahakoon et al. (2022a) to estimate parameters. This algorithm uses two steps. The first step consists of considering each outbreak independently and obtaining samples from posterior distributions of sub-population-specific parameters. In this step, any Approximate Bayesian Computation (ABC)-based algorithm can be used. We used the ABC-SMC algorithm of Toni, Welch, Strelkowa, Ipsen, and Stumpf (2009). The posterior samples are then used within an importance sampling context to construct an approximate likelihood at the hyper-parametric level. Using this likelihood, the posterior samples for the hyper-parameters are obtained within a pseudo-marginal setting. Finally, under the second step, the sub-population-specific parameters under a hierarchical model are obtained by sampling from the conditional prior given the estimated hyper-parameters. For a detailed explanation of the algorithm construction and related theory, refer to Alahakoon et al. (2022a). In relation to the application of the algorithm in this study, for an explanation of model calibration, and relevant diagnostics, see the Supplementary Material.

When considering all the sub-populations independently, we used weakly informative prior distributions on *β*_*j*_ and *γ*_*j*_. They were uniform (0.001,10) and uniform (0.00001, 3) respectively. Following the reference of Alahakoon et al. (2022b), we used a strong informative prior on the waning immunity rate, TN(0.03, 0.01^2^, 0.01, 0.2). Under a hierarchical framework, we used uniform (0.001, 10) and uniform (0, 2.5) as priors for *ψ*_*β*_ and *σ*_*β*_, uniform (0.00001, 3) and uniform (0,1) as priors for *ψ*_*γ*_ and *σ*_*γ*_, and TN(0.03, 0.01^2^, 0.01, 0.2) and uniform (0, 0.15) as priors for *ψ*_*μ*_ and *σ*_*μ*_.

Under the first scenario, we conducted parameter estimation under a hierarchical framework first using our knowledge of which group each sub-population is in. Under the second scenario, prior to the hierarchical analysis, we applied the k-means clustering algorithm to the posterior samples of the independently estimated transmission rates to identify clusters within the 30 sub-populations. We used posterior medians and 0.025% and 0.975% quantiles of the posterior samples as inputs to the k-means algorithm. See Supplementary Material S2.2 for an explanation of our usage of the k-means algorithm and selection of the optimal number of clusters. Next, given the identified sub-populations within each cluster, we conducted parameter estimation with a hierarchical modeling framework applied to each cluster (independently). In this setting, we did not introduce an additional (fourth) level into the hierarchical framework. We explore the potential to construct a hierarchical model with four levels in the Discussion.

## 4 Results

In this section, we present the results related to the transmission rates. The interested reader is referred to the Supplementary Material for visualisations and diagnostics related to recovery and waning immunity rates.

Figure 3 illustrates the marginal posterior distributions of hyper-parameters *ψ*_*β*_ and *σ*_*β*_ under the first scenario in which groups are known. Table 3 displays the posterior medians and 95% Highest Posterior Density (HPD) intervals. As expected, there is a notable difference between the posterior distributions for the hyper-mean for transmission, *ψ*_*β*_ for the three groups. The posterior distributions were located around the medians 3.48, 1.979, and 2.551 for Sub-groups A (high transmission), B (low transmission), and C (medium transmission) respectively with excellent correspondence to the true values (asterisks in Figure 3, and Table 2). When the hierarchical analysis was done by considering all the sub-populations together (one-group), the posterior distribution of the hyper-mean for transmission was located between the posterior distribution of high and low sub-groups with a posterior median of 2.608, and, as expected, the standard deviation notably increased, matching the target values (asterisks in Figure 3, and Table 2).

**Table 3:** Posterior medians and 95% Highest Posterior Density (HPD) intervals for Ψ_*β*_ and *σ*_*β*_ obtained by considering the true sub-groups

| Parameter | Sub-group A | Sub-group B | Sub-group C | One group |
| --- | --- | --- | --- | --- |
| $\Psi_\beta$ | 3.480<br>(3.246, 3.834) | 1.979<br>(1.836, 2.198) | 2.551<br>(2.219, 2.786) | 2.608<br>(2.399, 2.875) |
| $\sigma_\beta$ | 0.359<br>(0.010, 0.659) | 0.159<br>(0.003, 0.301) | 0.373<br>(0.182, 0.595) | 0.643<br>(0.449, 0.901) |

**Figure 3:**
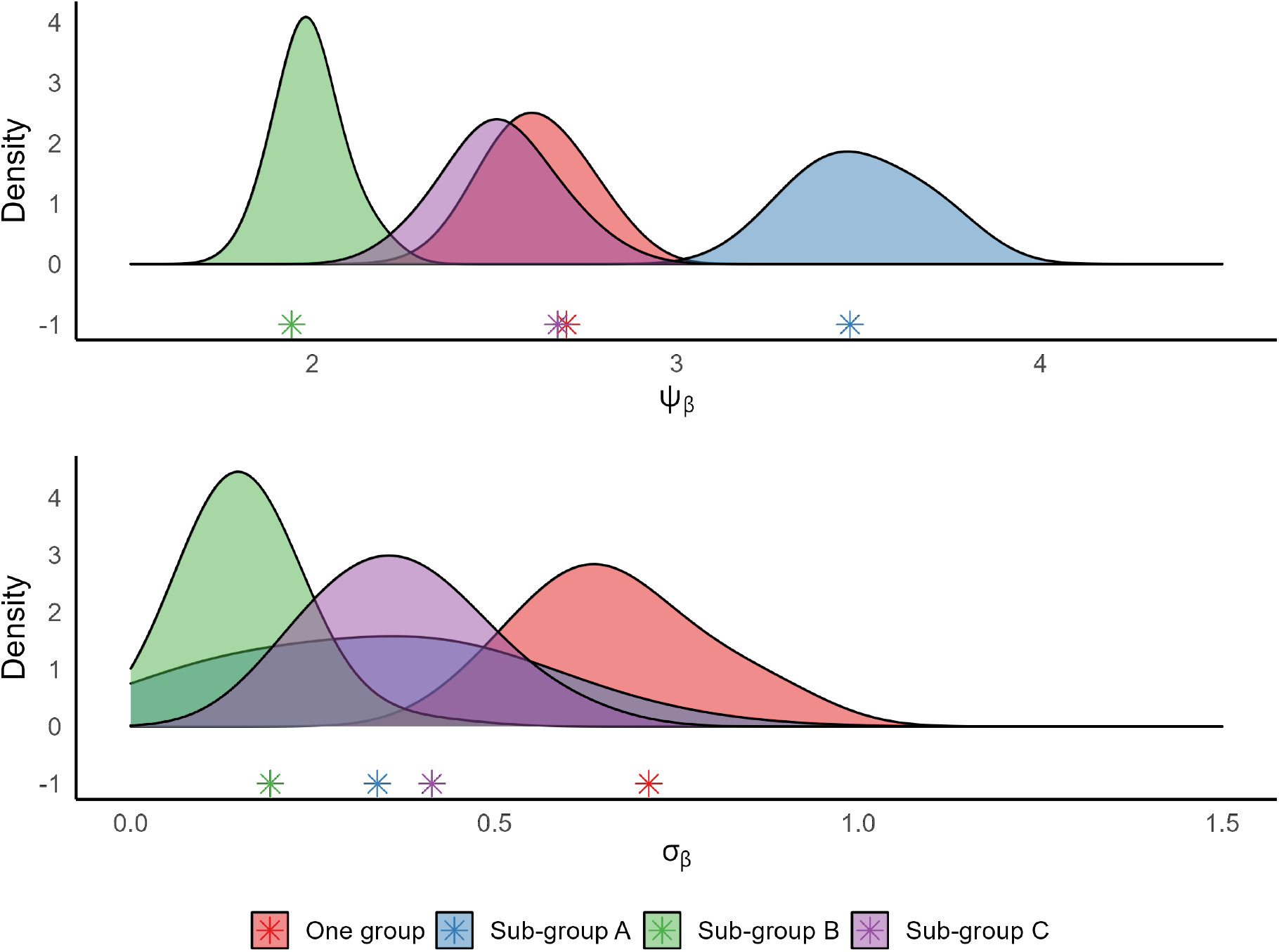
Posterior distributions for mean hyper-parameters with a hierarchical analysis considering each group and combining all three groups together. The asterisks are the group means of the parameters of each group.

See Supplementary Material S2.1 for the posterior distributions of *ψ*_*γ*_, *σ*_*γ*_, *ψ*_*μ*_, and *σ*_*μ*_. For *ψ*_*γ*_, *σ*_*γ*_, *ψ*_*μ*_, and *σ*_*μ*_, posterior medians for all three groups and that under the one-group analysis were similar.

In the second scenario, in which we do not assume the groupings are known, we first carried out the k-means analysis before conducting the hierarchical analysis. Figure 4 illustrates the results. The k-means analysis identified that the optimal number of clusters for the 30 sub-populations was two (see Supplementary Material S2.2). We also show the 3-cluster analysis for comparison, given we have 3 true groups. When two clusters were specified, we obtained 11 sub-populations for Cluster 1 and 19 sub-populations for Cluster 2. The majority of the sub-populations in Cluster 1 contained the sub-populations in Sub-group A and those in Cluster 2 contained the sub-populations in Sub-group B and C. Therefore for convenience, we refer to these as the Predicted High transmission group and Predicted Low/Medium transmission group respectively. We label these clusters as such for the purpose of further analysis and evaluation of the performance of the clustering algorithm. When three clusters were specified, Cluster 1 (Predicted High transmission) consisted of 8 sub-populations, Cluster 2 (Predicted Low transmission) consisted of 14 sub-populations, and Cluster 3 (Predicted Medium transmission) consisted of 8 sub-populations.

**Figure 4:**
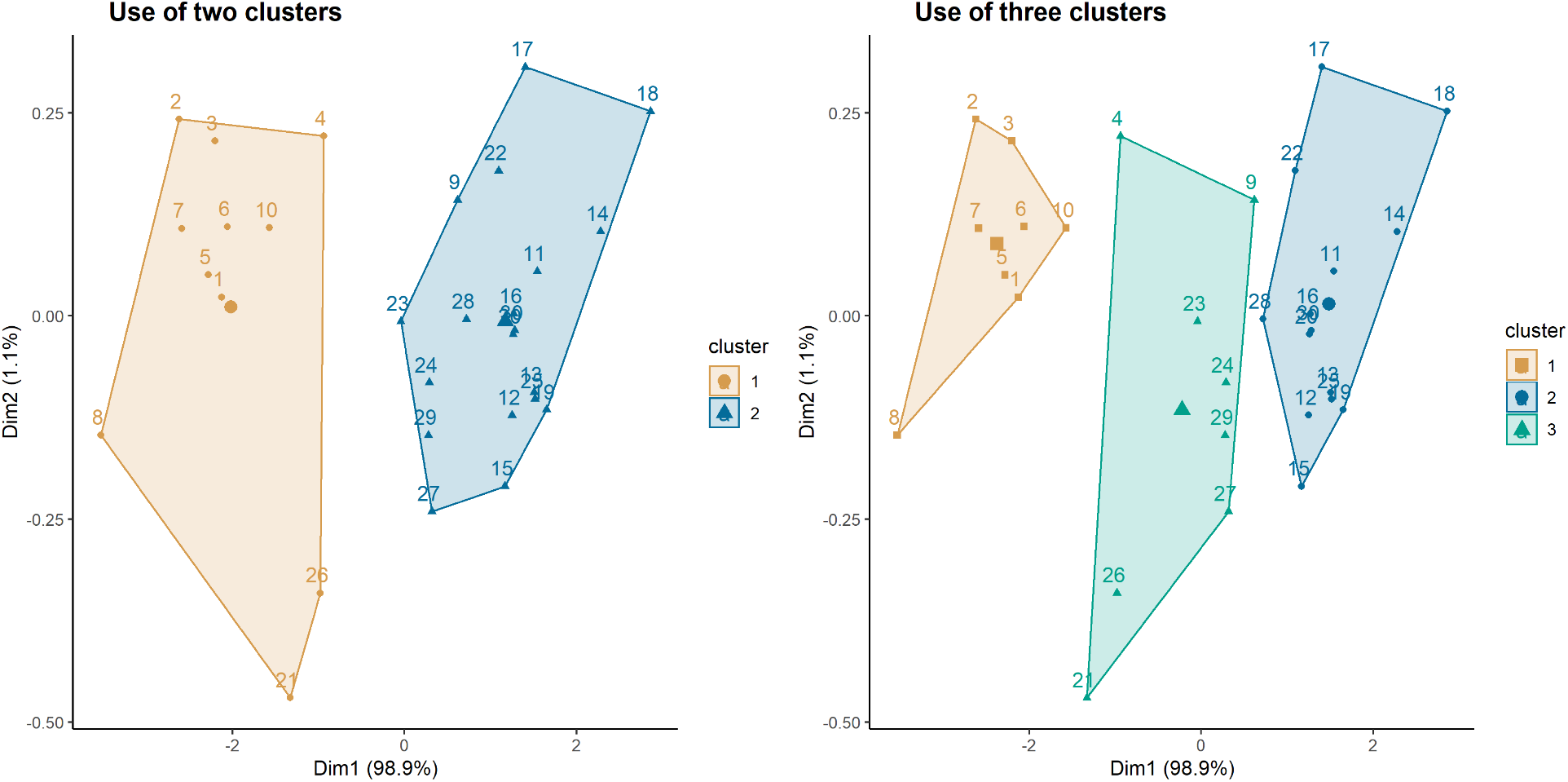
Clusters generated from the k-means algorithm.

Having identified the sub-populations included in the (2 or 3) clusters, we conducted a hierarchical analysis for each cluster. Figure 5 illustrates the posterior distributions of *ψ*_*β*_ for two and three clusters respectively. Table 4 presents the posterior medians and 95% HPD intervals. A comparison is done with the posterior distribution of *ψ*_*β*_ with no clustering (all 30 sub-populations). The posterior medians of *ψ*_*β*_ under two clusters were 3.502 for high and 2.155 for low/medium transmission levels, and those under three clusters were 3.720, 2.029, and 2.786 for high, low, and medium transmission levels respectively. See Supplementary Material S2.2 for a visualisation of the posterior distributions of hyper-parameters for recovery and waning immunity.

**Table 4:** Posterior medians and 95% Highest Posterior Density (HPD) intervals of Ψ_*β*_ after cluster analysis

| Two clusters | Parameter | Cluster 1 | Cluster 2 |  |
| --- | --- | --- | --- | --- |
| | $\Psi_\beta$ | 3.502<br>(3.267, 3.769) | 2.155<br>(2.026, 2.287) | |
| Three clusters | Parameter | Cluster 1 | Cluster 2 | Cluster 3 |
| | $\Psi_\beta$ | 3.720<br>(3.410, 4.135) | 2.029<br>(1.876, 2.152) | 2.786<br>(2.525, 3.087) |

**Figure 5:**
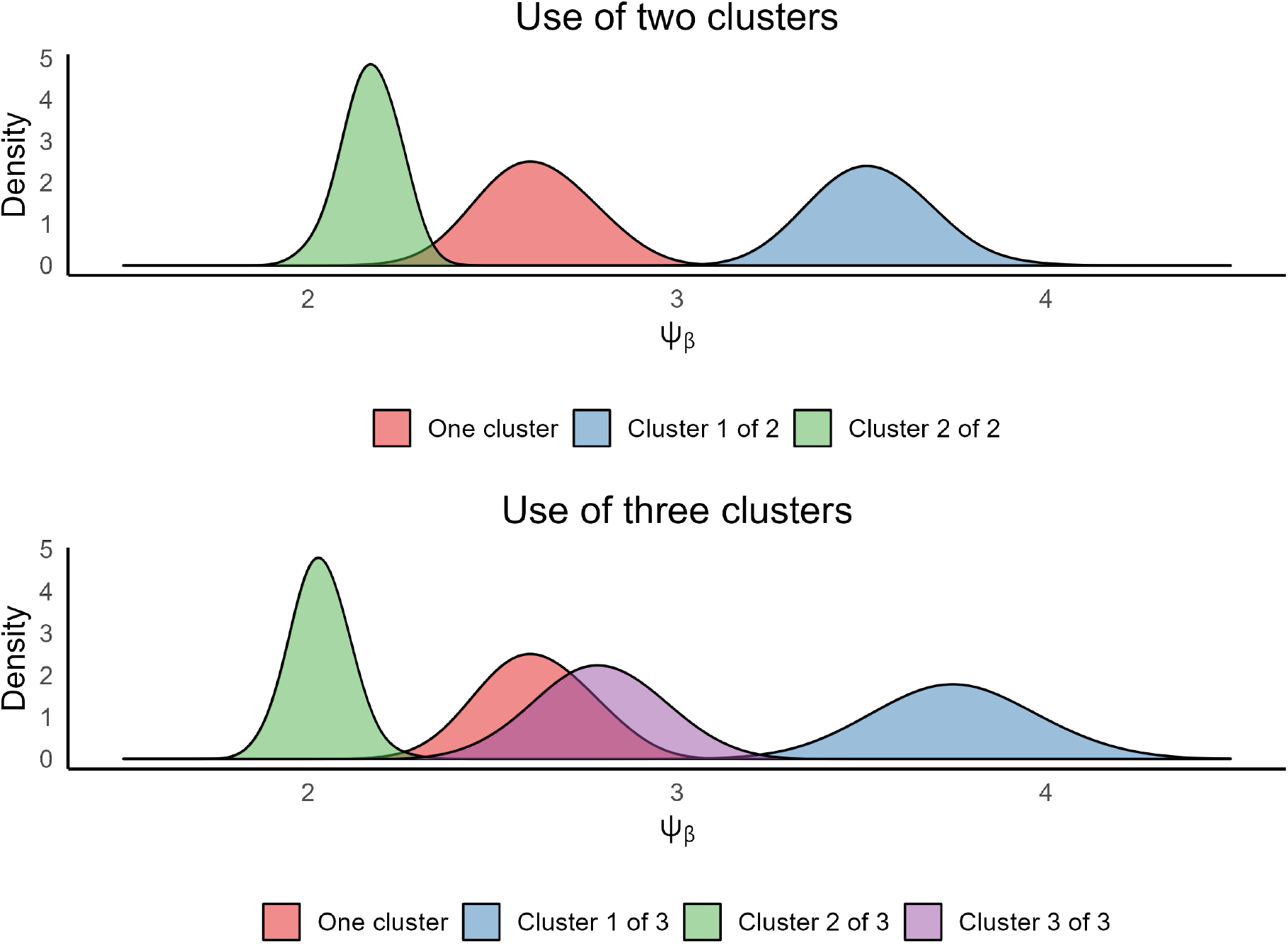
Posterior distributions for mean hyper-parameters with a hierarchical analysis applied to each cluster (2 clusters in the top figure and 3 clusters in the bottom figure). A reference analysis without clustering (one-cluster) is also shown.

Figure 6 illustrates the sub-population specific parameters estimated under hierarchical estimation after identifying the subsets of sub-populations by clustering. A visual comparison can be made with the corresponding posterior distributions estimated under the assumption of all 30 sub-populations belonging to a single cluster (posterior distributions in grey). When two clusters were used, the posterior distributions of the sub-populations in Cluster 1 (the Predicted High transmission group) shifted towards the hyper-mean of Cluster 1. Similarly, the posterior distributions of the sub-populations in Cluster 2 (the Predicted Low/Medium transmission group) shifted towards the hyper-mean of Cluster 2. On the other hand, when no clustering was used, as expected, the posterior distributions of all the sub-populations were pulled towards the hyper-mean of the single group. Furthermore, compared to the posterior distributions with clustering, these posterior distributions had a higher variability. We observe similar behaviour when three clusters were used.

**Figure 6:**
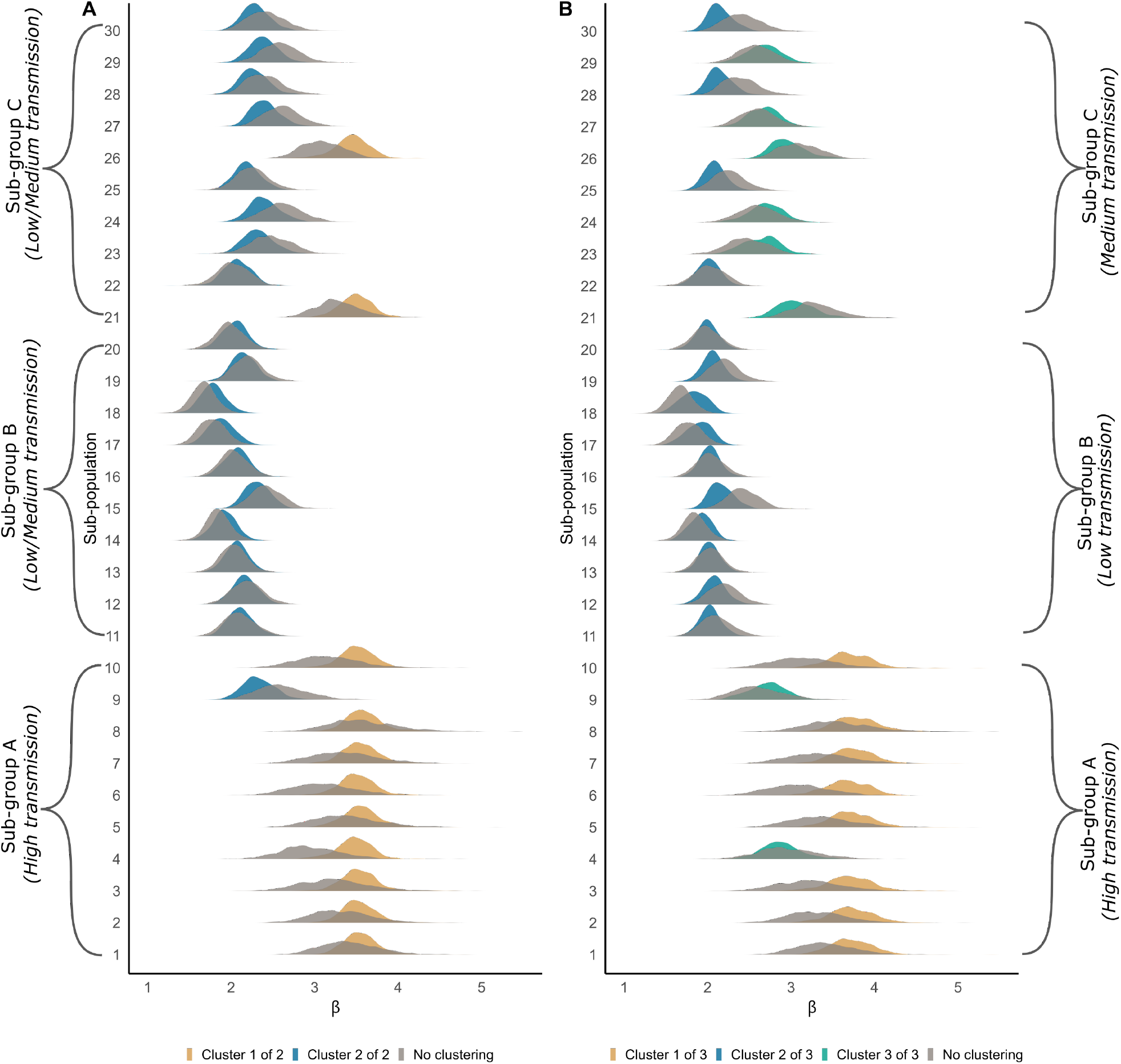
Posterior distributions for transmission rates under a hierarchical analysis for sub-populations.

When two clusters were used, sub-population 9, which is from Sub-group A (High transmission), was incorrectly allocated to the Low/Medium transmission cluster. When three clusters were used, both sub-populations 4 and 9 from Sub-group A, were incorrectly clustered. This can be understood by noting that the sampled value of *β* for sub-populations 4 and 9 were 3.091 and 2.981 respectively, substantially different from all the other high transmission parameter values in Sub-group A.

To identify how accurate our clustering was, we calculated the confusion matrix using the *caret* package in R. Tables 5 and 6 illustrate the results. For two clusters, when the transmission rate was high, nine out of ten sub-populations classified the transmission rate as high whereas when the transmission was low/medium, two out of twenty sub-populations were predicted as having high transmission (Table 5). The sensitivity (the proportion of true high transmissions) was 0.9 and the specificity (the proportion of true low/medium transmission) was 0.9. Overall, only 3 sub-populations were incorrectly clustered (that is, the accuracy was 0.9).

**Table 5:** Summary statistics related to using two clusters

|  | True High | True Low/Medium |
| --- | --- | --- |
| Predicted High | 9 | 2 |
| Predicted Low/Medium | 1 | 18 |
| Sensitivity = | 0.9 |  |
| Specificity = | 0.9 |  |
| Accuracy = | 0.9 |  |

**Table 6:** Summary statistics related to using three clusters

|  | True High | True Low | True Medium |
| --- | --- | --- | --- |
| Predicted High | 8 | 0 | 0 |
| Predicted Low | 0 | 10 | 4 |
| Predicted Medium | 2 | 0 | 6 |
| Sensitivity (by class) | 0.8 | 1.0 | 0.6 |
| Specificity (by class) | 1.0 | 0.8 | 0.9 |
| Accuracy = | 0.8 |  |  |

Alternatively, if one were to label Cluster 1 as Predicted High/Medium and Cluster 2 as Predicted Low, then the sensitivity (the proportion of true High/Medium) was 0.45, specificity (the proportion of true Low) was 0.8, and the accuracy was 0.567. While labelling the two clusters in this way is not incorrect, our choice to label Cluster 1 as Predicted High and Cluster 2 as Predicted Low/Medium was pragmatic as our data are synthetic, and we had perfect knowledge about the true grouping of the sub-populations.

Table 6 shows corresponding results when using three clusters. Two sub-populations were predicted as having medium transmission when they were actually from the high transmission group. All the low transmission sub-populations were correctly identified. Four out of ten sub-populations that had medium transmission rates were allocated to the low transmission cluster. The accuracy for using three clusters was 0.8. Overall accuracy, specificity, and sensitivity were higher for the two cluster analysis.

### 4.1 Assessing the parameter estimates of sub-populations under different estimation regimes

In order to assess if the estimates for the transmission rates of the sub-populations improved when subsets of sub-populations were identified through cluster analysis, we used the Region of Practical Equivalence (ROPE) criteria (J. Kruschke, 2014; J. K. Kruschke, 2013) along with the 95% HPD intervals (Chen, Shao, & Ibrahim, 2012) to assess the posterior distributions under independent estimation and hierarchical estimation. For each sub-population, we defined a ROPE interval around its true parameter value (*β*_*j*_ *± c*) where *c* is a pre-defined value. Then we calculated the 95% HPD intervals for all the sub-populations under the different estimation methods we used. Next, we calculated the percentage of the HPD intervals that fall within the ROPE interval (See Figure 7 for an illustration for sub-population 1 with *c* = 0.5). We used the *BEST* package in R (J. K. Kruschke & Meredith, 2020) to compute these values.

**Figure 7:**
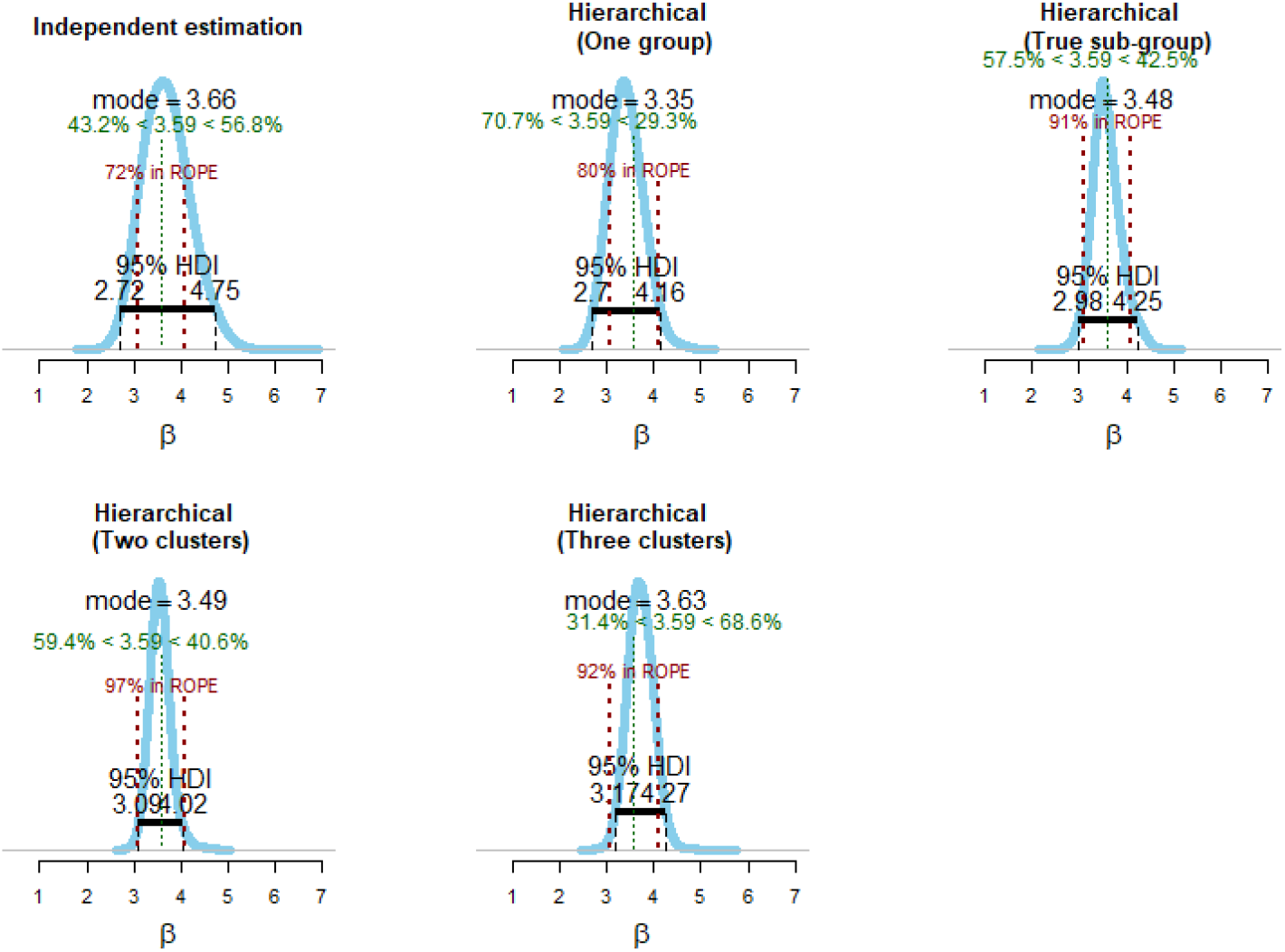
A comparison of 95% HPD intervals and ROPE intervals under different estimation methods for sub-population 1.

Figure 8 shows the distributions of the percentages over the different sub-populations when *c* = 0.5. Overall performance was highest under the 2-cluster hierarchical analysis, with the distribution of ROPE percentage coverage marginally higher than even under the hierarchical analysis conducted on the three true groups. Furthermore, for this set of 30 sub-populations, independent parameter estimation under-performed in comparison to hierarchical estimation methods. Supplementary Material S4 presents the ROPE analysis under different values of *c*.

**Figure 8:**
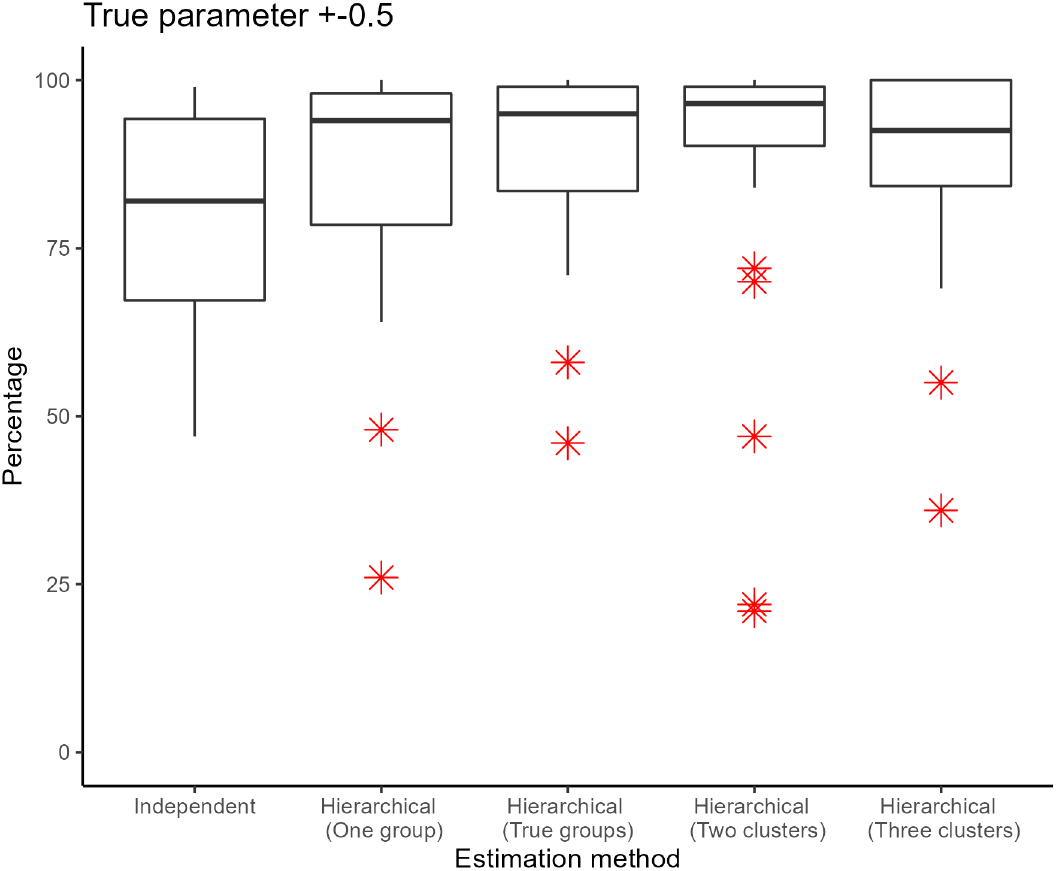
Box plots of the percentage of the 95% HPD intervals that intersect the ROPE intervals taken as observations across the 30 sub-populations under different estimation methods.

However, as expected (due to shrinkage), not all sub-population specific estimates performed well under the hierarchical methods. Figure 9 shows box plots of the percentages under the different estimation methods in comparison to independent estimation with ties (light grey lines linking sub-populations). Generally, parameter estimates improved with hierarchical estimation except for a few sub-populations (notice the upward trend in the lines). For most parameter estimates that did not perform well under hierarchical methods, the estimates under independent estimation also did not perform particularly well (notice the lower quartile of the box plots).

**Figure 9:**
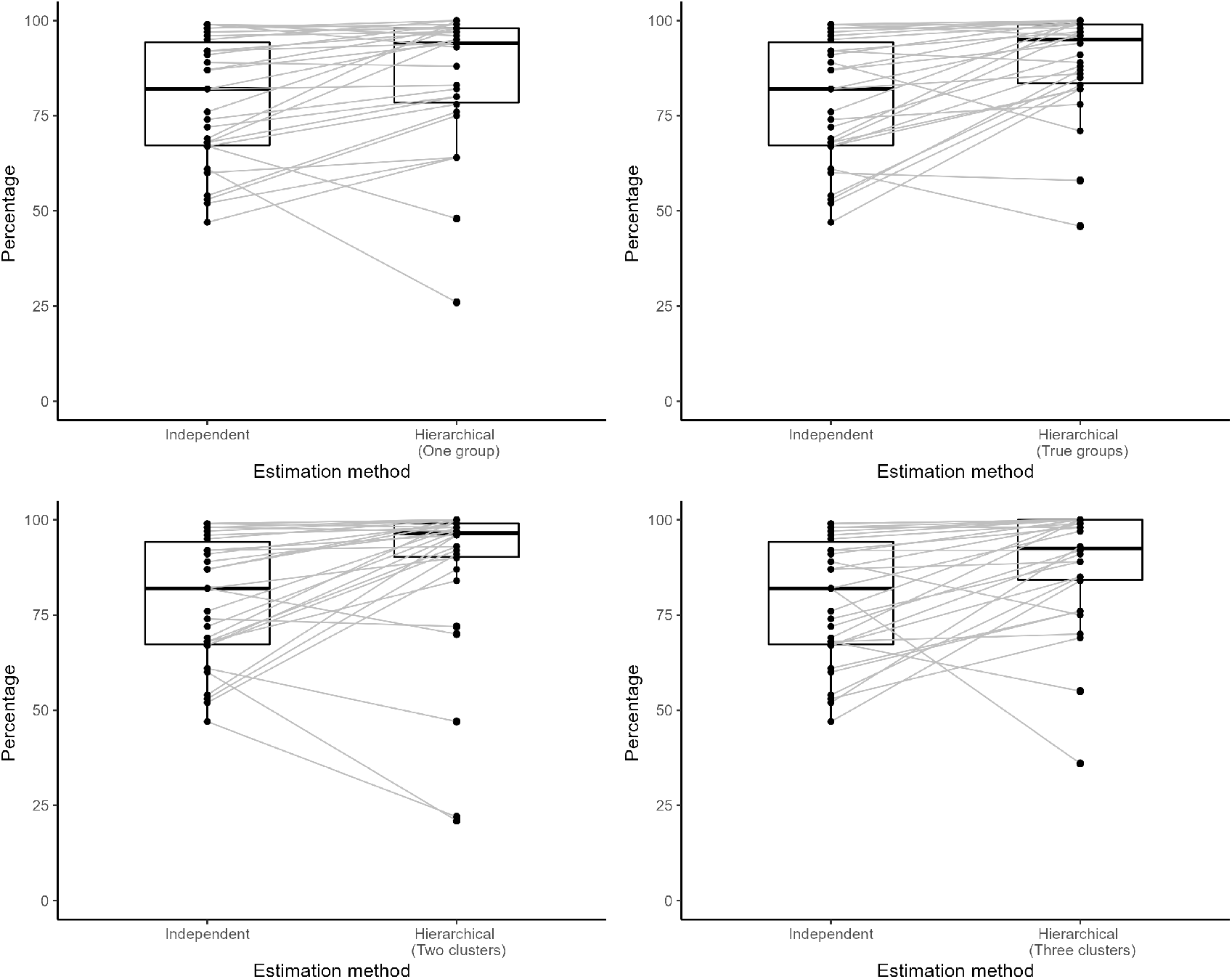
Box plots of the percentage of the 95% HPD intervals that intersect the ROPE intervals taken as observations across the 30 sub-populations under different estimation methods. Grey lines are corresponding sub-populations under each estimation method.

## 5 Discussion

Although hierarchical models generally improve parameter estimates at the sub-population level, due to induced shrinkage towards a common mean, there is the possibility for extreme parameter values to be over or underestimated. This common criticism of hierarchical models has been well described in the literature in various fields (McGlothlin & Viele, 2018; Scheibehenne & Pachur, 2013).

In the infectious disease epidemiology context, when there are a large number of outbreaks, there is a high possibility that the variability of the sub-population-specific parameters will be high due to the sparse characteristics of the sub-populations. Therefore, when a hierarchical analysis is carried out by considering all the sub-populations as one group, the estimated standard deviation of the conditional prior distribution may be high and the estimated mean of the conditional prior distribution will also reflect all the sub-population specific parameters. We have shown that in such situations, as expected, the posterior distributions of the sub-population-specific parameters may be wider and biased.

However, we have shown that this issue can be mitigated by identifying subsets of similar parameters (that is, subsets of parameters with less variability) from a preliminary non-hierarchical analysis. We have shown that the use of a common clustering algorithm such as the k-means algorithm can be used to identify clusters of sub-populations that had similar parameter estimates. Once an optimal number of clusters were identified, a hierarchical analysis on each subset yielded more accurate estimates for the sub-population specific parameters.

Unsurprisingly, the optimal number of clusters inferred from the preliminary analysis may not reflect the true sub-group number. In our dataset, sub-populations in Sub-group C had a high standard deviation among the transmission rates (0.5 compared to 0.25 and 0.15 of Sub-groups A and B), and the transmission rate parameter *β* was often close to that of either Sub-group A or B. Therefore, transmission rates for Sub-group C were at times grouped together with those of Sub-groups A or B. Nonetheless, once the optimal number of clusters was used to estimate the model parameters under a hierarchical analysis, the sub-population specific parameters improved compared to when three clusters were used.

While a clustering analysis improves parameter estimates, it may be of interest to study all the sub-populations within a hierarchical model with more than three levels. For instance, Level II may consist of conditional priors of clusters, Level III may consist of the conditional prior of all the sub-populations, and Level IV, the hyper-prior distributions. In our study, one hypothesis for the surprising result that two clusters outperformed three (clusters and groups) may be that the hierarchical model performed better with more sub-populations in each analysis. This may be overcome with a 4-level analysis as then all 30 sub-populations will share information.

We further note that the use of the additional clustering step did not substantially increase the overall computational cost as we employed the two-step algorithm of Alahakoon et al. (2022a). This is due to the fact that we re-used the posterior samples of the sub-population specific parameters obtained during the first step of the algorithm for the purpose of cluster analysis. However, we note that greater efficiency may be achieved by embedding a clustering step within the two-step algorithm. We will consider this in the future.

Another possible application of this framework lies within surveillance frameworks where a large number of outbreaks each evolving in isolation from the others are observed in communities with diverse characteristics. Although conducting a hierarchical analysis is generally favourable, identifying sub-groups of communities with similar model parameters prior to a hierarchical analysis may provide improved estimates of parameters. This in turn may be valuable for developing effective public health policy and intervention measures to suit different community groups.

Our estimation framework can also be applied outside an epidemiological context. It maybe applied to biological and ecological data where dynamics within sub-populations are expected to or identified to have distinct groupings.

This study had only considered a stochastic *SIRS* model structure and identifying subsets of sub-populations based on the transmission level only. It maybe of interest to apply this framework to other complex epidemic models and to identify subsets of sub-populations based on multiple parameters.

## Data Availability

All data produced are available online at https://github.com/PunyaAlahakoon/Use_of_Clusetring_in_hierarchical_models.git

## 6 Acknowledgements

Unless otherwise mentioned, computations were done in MATLAB or R across 32 clusters. All the computations were carried out by the use of the Nectar Research Cloud (project Infectious Diseases), a collaborative Australian research platform supported by the National Collaborative Research Infrastructure Strategy (NCRIS). All the plots were generated with ggplot2 (Wickham, 2016) in R. The codes are publicly available (see Supplementary Material for details).

## 7 Funding

Punya Alahakoon is supported by a Melbourne Research Scholarship from the University of Melbourne. P.G. Taylor would like to acknowledge the support of the Australian Research Council via the Centre of Excellence for Mathematical and Statistical Frontiers (ACEMS).

## Notes

### Competing Interest Statement

The authors have declared no competing interest.

