## Supplementary material for "Use of clustering to improve estimation of epidemic model parameters under a Bayesian hierarchical framework": Supplemenary_material

### S1 Independent parameter estimation

For parameter estimation by considering each outbreak independently, we used the ABC-SMC algorithm of Toni, Welch, Strelkowa, Ipsen, and Stumpf (2009). We pre-defined the tolerance values (see Table S1) across 7 generations. Our choice for choosing appropriate tolerance values was similar to that of Alahakoon, McCaw, and Taylor (2022a).

*

Table S1: Tolerance values across 7 generations for ANC-SMC algorithm

| | **Generation** | | | | | | |
|---|---|---|---|---|---|---|---|
| **Sub-population** | **1** | **2** | **3** | **4** | **5** | **6** | **7** |
| 1 | 282 | 250 | 230 | 180 | 150 | 110 | 70 |
| 2 | 239 | 219 | 180 | 150 | 120 | 100 | 65 |
| 3 | 247 | 220 | 200 | 180 | 150 | 100 | 95 |
| 4 | 235 | 220 | 200 | 180 | 150 | 100 | 70 |
| 5 | 266 | 230 | 200 | 180 | 150 | 100 | 90 |
| 6 | 211 | 200 | 180 | 150 | 125 | 90 | 65 |
| 7 | 266 | 250 | 200 | 180 | 150 | 120 | 90 |
| 8 | 265 | 250 | 220 | 200 | 150 | 100 | 65 |
| 9 | 219 | 200 | 180 | 150 | 130 | 100 | 60 |
| 10 | 261 | 250 | 200 | 180 | 150 | 120 | 95 |
| 11 | 315 | 300 | 250 | 200 | 180 | 85 | 70 |
| 12 | 331 | 350 | 250 | 200 | 180 | 80 | 70 |
| 13 | 292 | 200 | 180 | 150 | 100 | 50 | 45 |
| 14 | 275 | 230 | 200 | 180 | 125 | 85 | 60 |
| 15 | 396 | 250 | 200 | 150 | 110 | 80 | 60 |
| 16 | 292 | 380 | 330 | 260 | 200 | 100 | 65 |
| 17 | 229 | 360 | 300 | 250 | 155 | 100 | 80 |
| 18 | 223 | 280 | 250 | 200 | 160 | 100 | 45 |
| 19 | 392 | 300 | 250 | 200 | 150 | 120 | 90 |
| 20 | 271 | 330 | 280 | 230 | 180 | 100 | 60 |
| 21 | 562 | 500 | 450 | 400 | 300 | 200 | 140 |
| 22 | 273 | 250 | 220 | 200 | 180 | 140 | 100 |
| 23 | 345 | 300 | 250 | 200 | 180 | 130 | 90 |
| 24 | 397 | 300 | 260 | 230 | 200 | 150 | 90 |
| 25 | 373 | 350 | 300 | 250 | 200 | 150 | 90 |
| 26 | 489 | 450 | 400 | 350 | 250 | 200 | 100 |
| 27 | 420 | 380 | 300 | 250 | 240 | 170 | 85 |
| 28 | 369 | 300 | 270 | 230 | 200 | 150 | 100 |
| 29 | 425 | 350 | 300 | 250 | 200 | 150 | 100 |
| 30 | 411 | 380 | 350 | 300 | 250 | 200 | 95 |

Figures S1 , S2, S3 show the marginal posterior distributions of parameters $(\beta, \gamma, \mu)$ of the 30 sub-populations. The relevant HPD intervals (calculated from *HDInterval* package of R) and the true parameter values are included as files in GitHub (see link below).

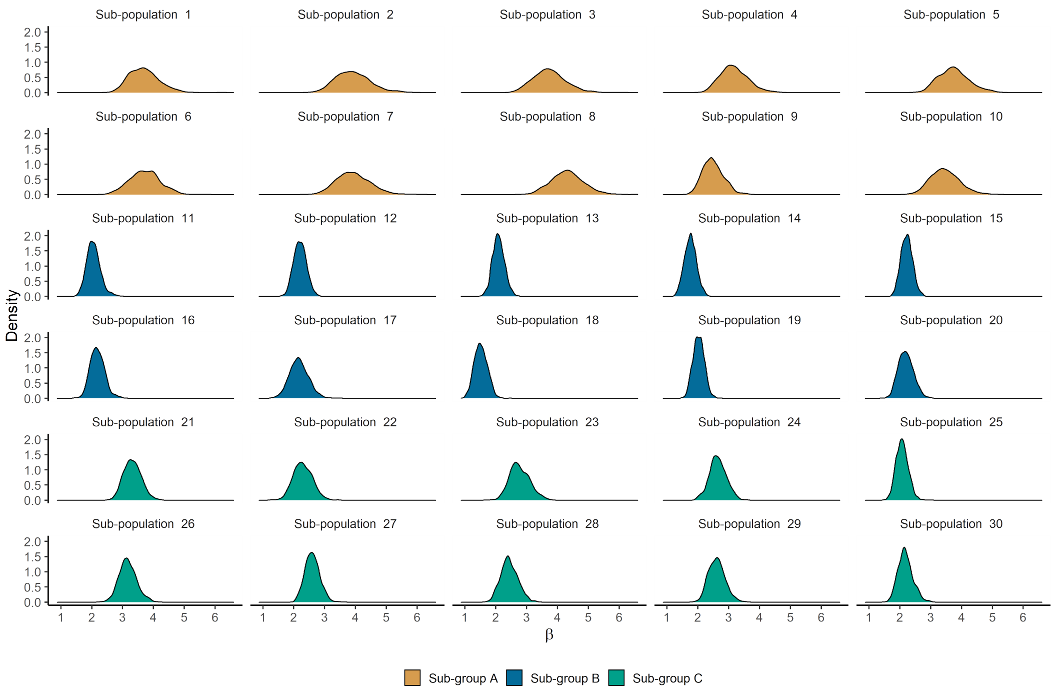


Figure S1: Marginal posterior distributions of $\beta_k, k = 1, 2, \ldots, 30$

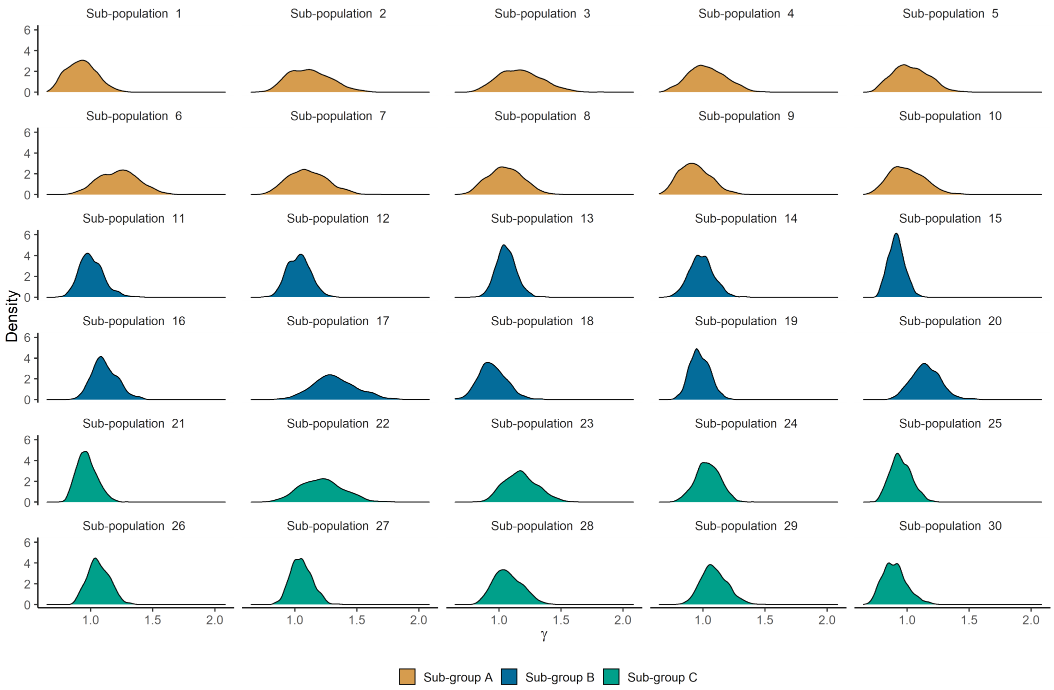


Figure S2: Marginal posterior distributions of $\gamma_k, k = 1, 2, \ldots, 30$

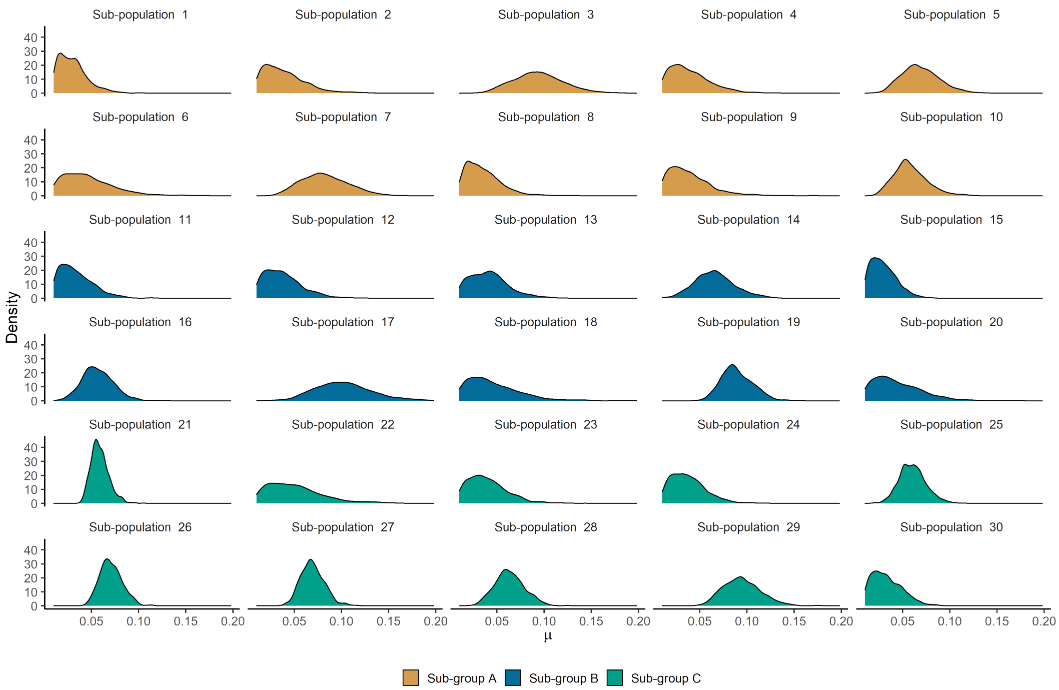


Figure S3: Marginal posterior distributions of $\mu_k, k = 1, 2, \ldots, 30$

### S2 Hierarchical parameter estimation

We used the two-step algorithm of (Alahakoon et al., 2022a) to estimate parameters of a stochastic hierarchical model.

#### S2.1 Hierarchical estimation by assuming that the sub-group is known

We generated $\beta$s, $\gamma$s, and $\mu$s from independent truncated normal distributions. Therefore, we constructed the likelihood function at the hyper-parametric level with independent multivariate normal distributions with no correlation between the hyper-parameters. See Alahakoon, McCaw, and Taylor (2022b) for a detailed explanation of this. The approximated likelihood at the hyper-parametric level is

$$\hat{p}(\boldsymbol{y}|\boldsymbol{\Psi}) \propto \prod_{k=1}^{15} \sum_{j=1}^{N_1} \frac{p(\boldsymbol{\theta}_k^{(j)}|\boldsymbol{\Psi})}{p(\boldsymbol{\theta}_k^{(j)})}, \tag{S.1}$$

The estimation procedure is similar to that of Alahakoon et al. (2022b). We direct the reader to Alahakoon et al. (2022b) for the algorithm and pseudo-code.

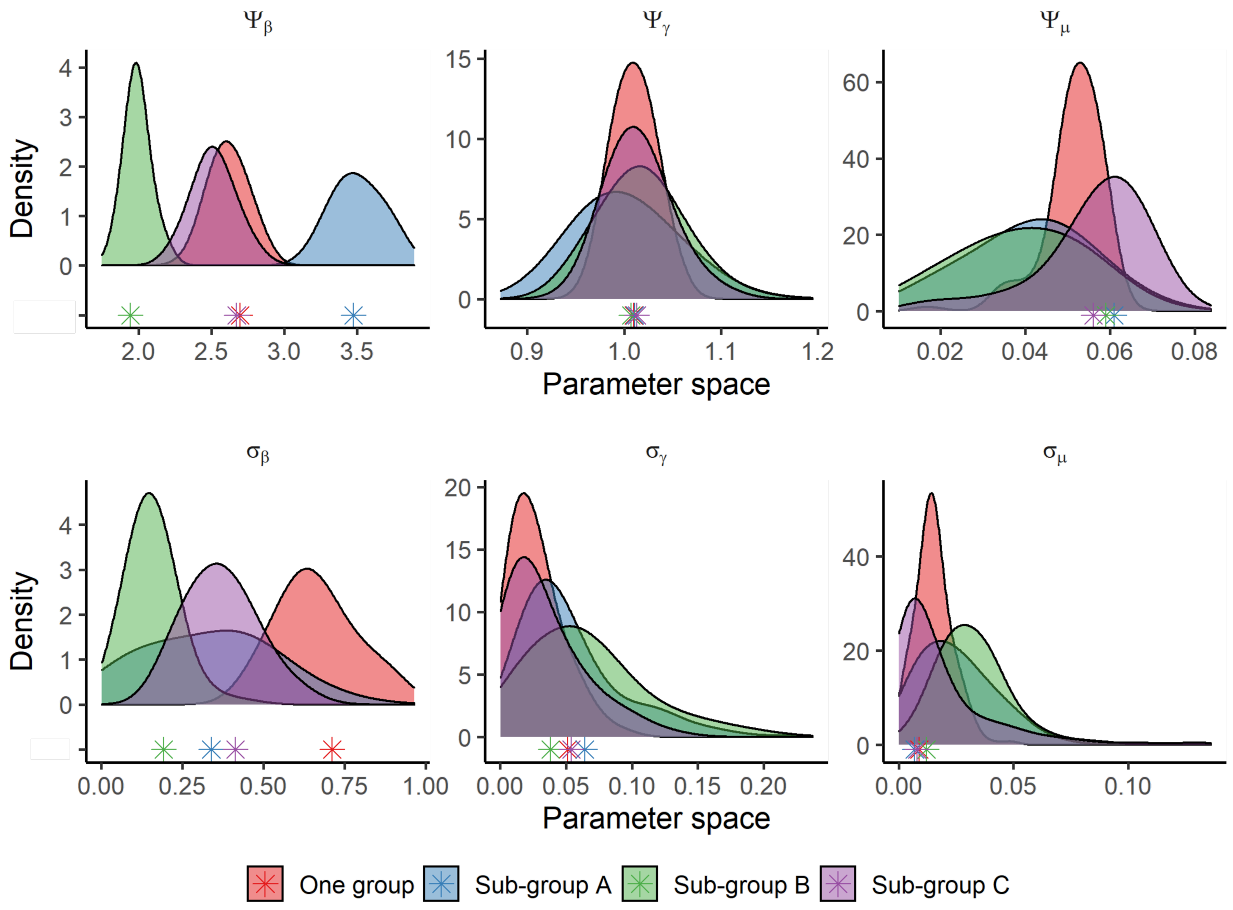


Figure S4: Marginal posterior distributions of hyper-parameters.

### S2.2 Hierarchical estimation by assuming that the sub-group is unknown

We carried out cluster analysis using the $k$-means algorithm. We used *factoextra* package in R to carry out the analysis. For each sub-population, we used the posterior samples of $\beta$ by considering each outbreak independently. We used the posterior median, 95% th quantile as data for clustering.

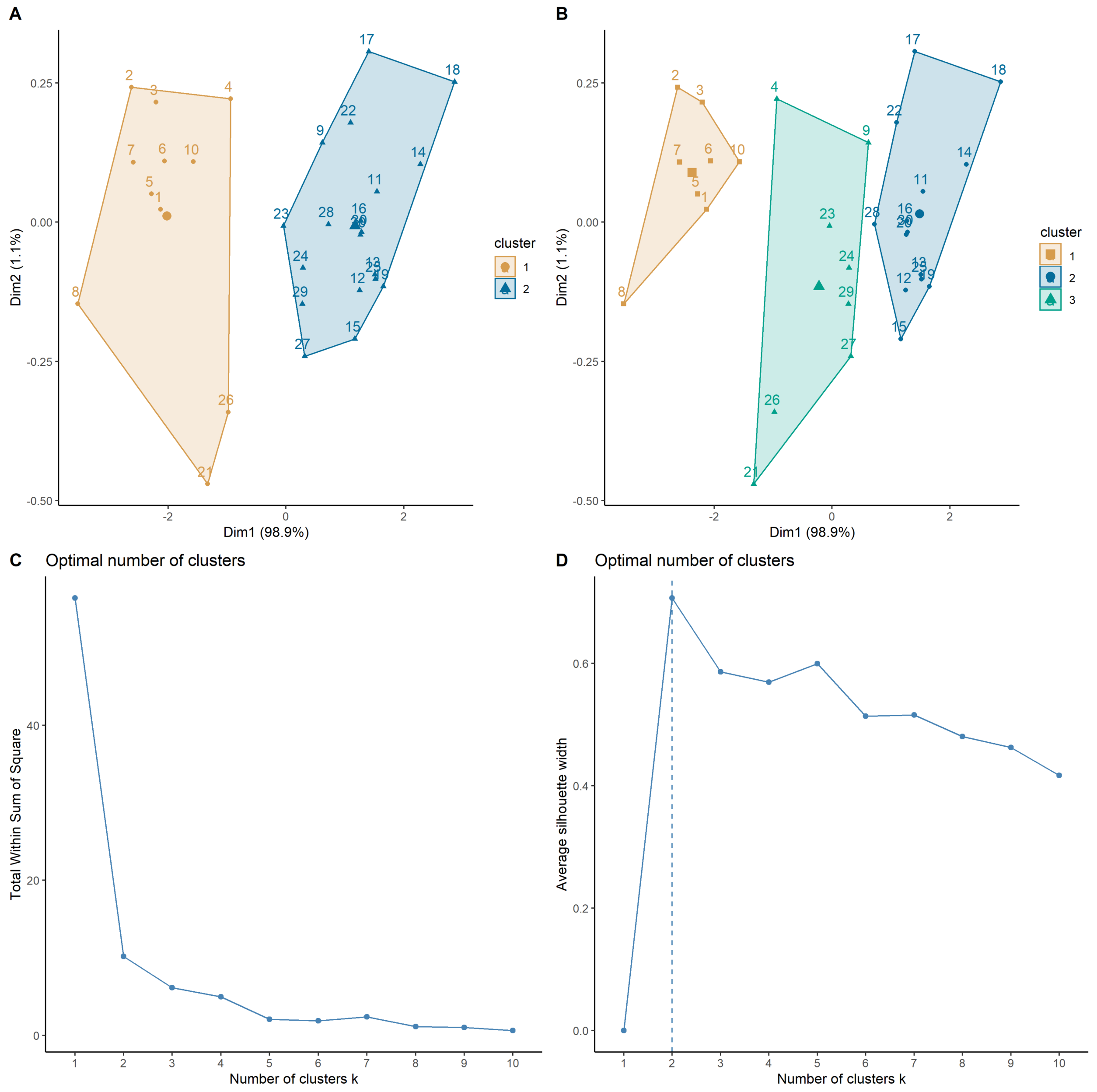


Figure S5: **Panel A**: Clustering with two clusters. **Panel B**: Clustering with three clusters. **Panel C**: Optimal number of clusters using total within the sum of squares. (Also known as the elbow method. The optimal cluster is the location of the bend) **Panel C**: Optimal number of clusters based on average silhouette width criterion (the highest average silhouette width indicates the optimal clustering number).

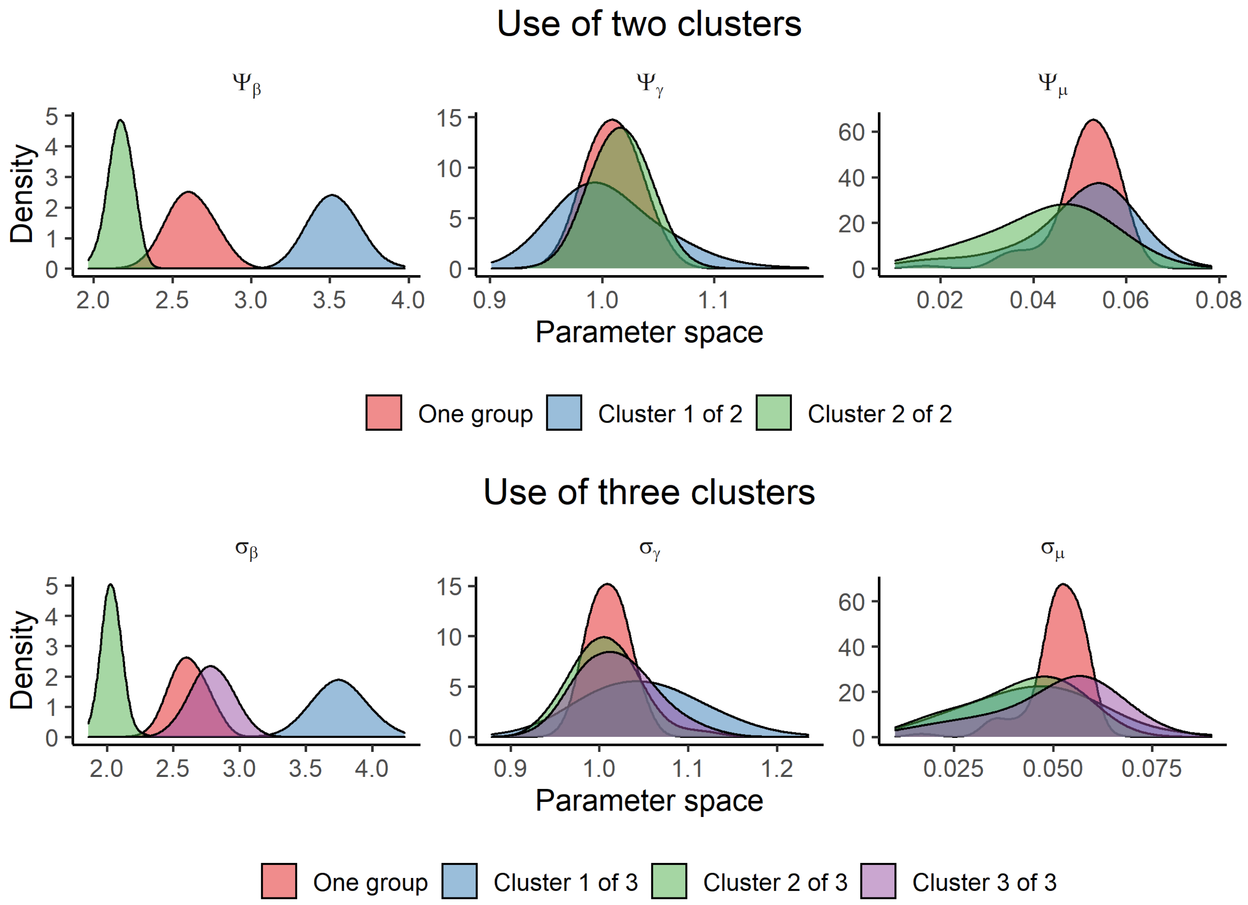


Figure S6: Marginal posterior distributions of the hyper-parameters after clustering.

### S3 Estimation of sub-population specific parameters

Under Step 2 of the two-step algorithm, we used a basic ABC algorithm and sampled from the conditional prior, $p(\boldsymbol{\theta}_k|\Psi)$ as in the algorithm of Alahakoon et al. (2022a). As tolerance levels, we used the tolerance levels of 7th generation in the ABC-SMC algorithm for the sub-populations.

### S3.1 Estimation by assuming that the sub-group is known

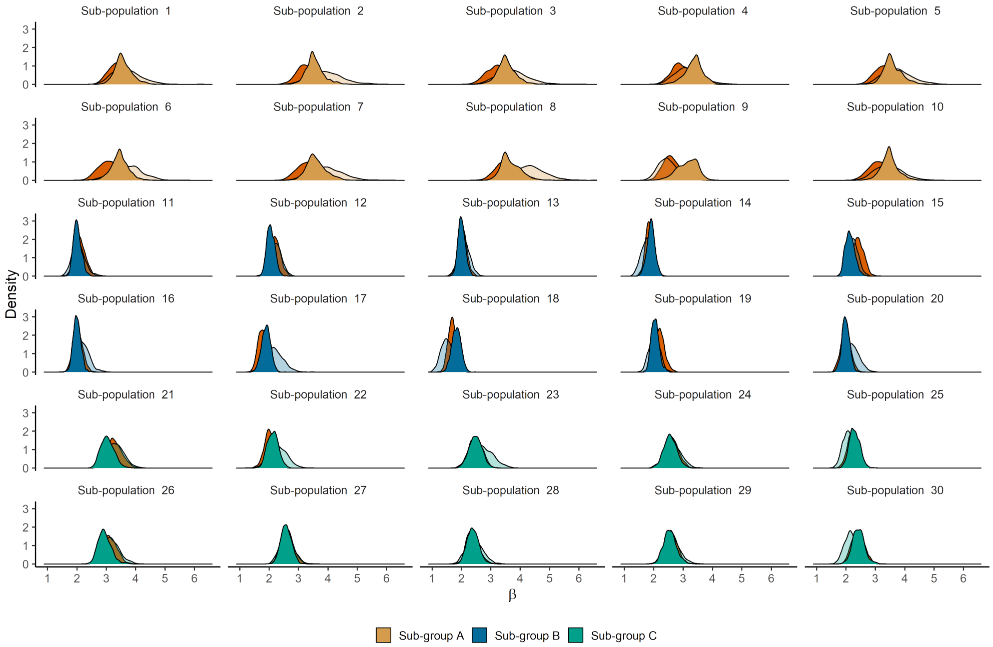


Figure S7: Marginal posterior distributions of $\beta_k, k = 1, 2, \ldots, 30$. **Dark shades:** hierarchical estimation done using the sub-populations of the true sub-group. **Light shades:** Parameter estimation is done by considering each sub-population independently. **Dark orange:** Hierarchical estimation done by considering all the sub-populations as one group.

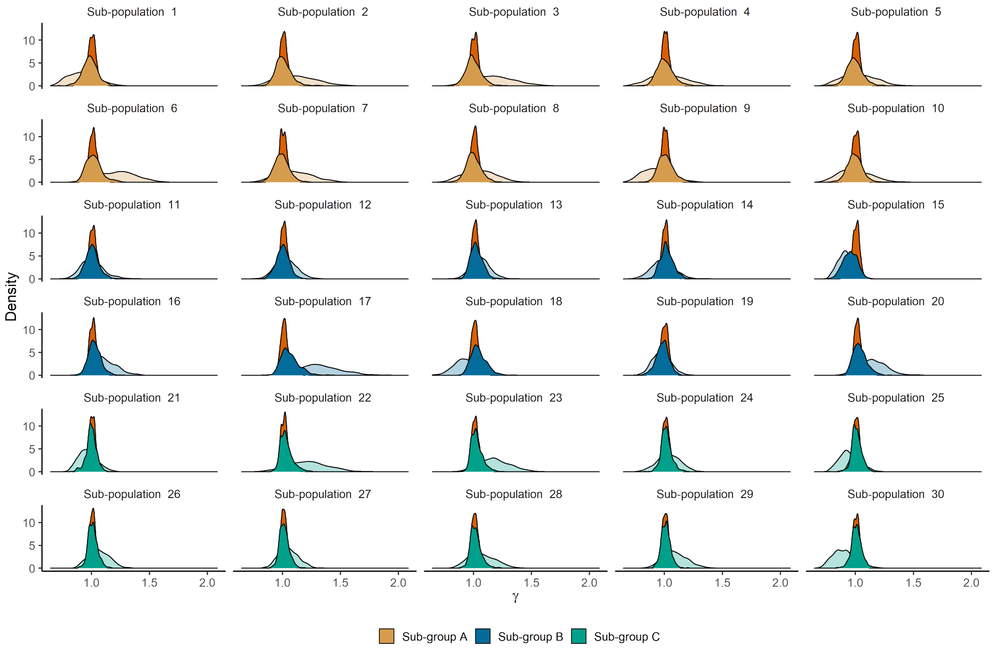


Figure S8: Marginal posterior distributions of $\gamma_k, k = 1, 2, \ldots, 30$. **Dark shades:** hierarchical estimation done using the sub-populations of the true sub-group. **Light shades:** Parameter estimation is done by considering each sub-population independently. **Dark orange:** Hierarchical estimation done by considering all the sub-populations as one group.

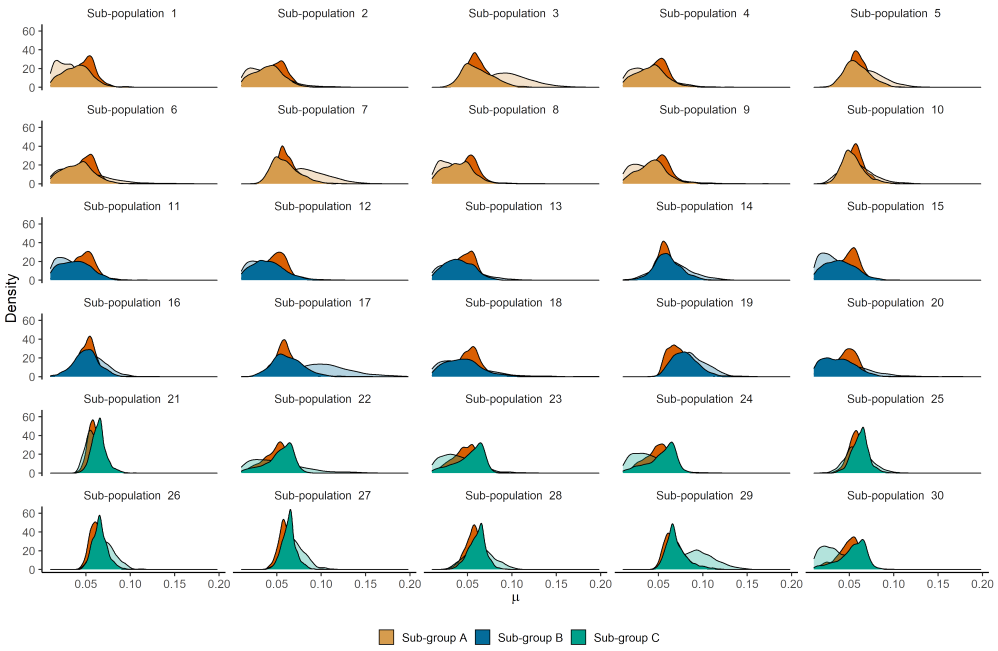


Figure S9: Marginal posterior distributions of $\mu_k, k = 1, 2, \ldots, 30$. **Dark shades:** hierarchical estimation done using the sub-populations of the true sub-group. **Light shades:** Parameter estimation is done by considering each sub-population independently. **Dark orange:** Hierarchical estimation done by considering all the sub-populations as one group.

#### S3.2 Estimation by assuming that the sub-group is unknown

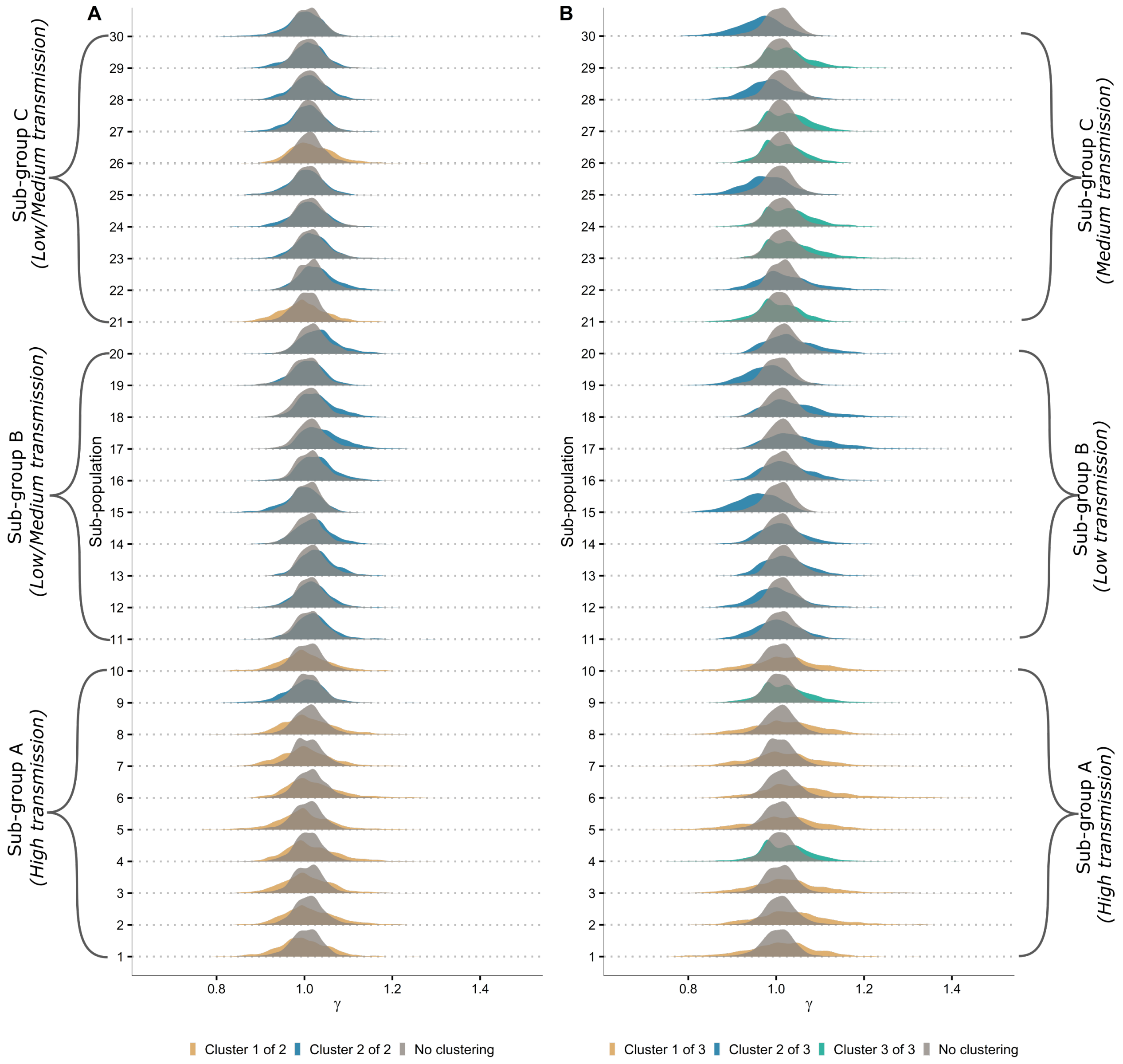


Figure S10: Marginal posterior distributions of $\gamma_k, k = 1, 2, \ldots, 30$

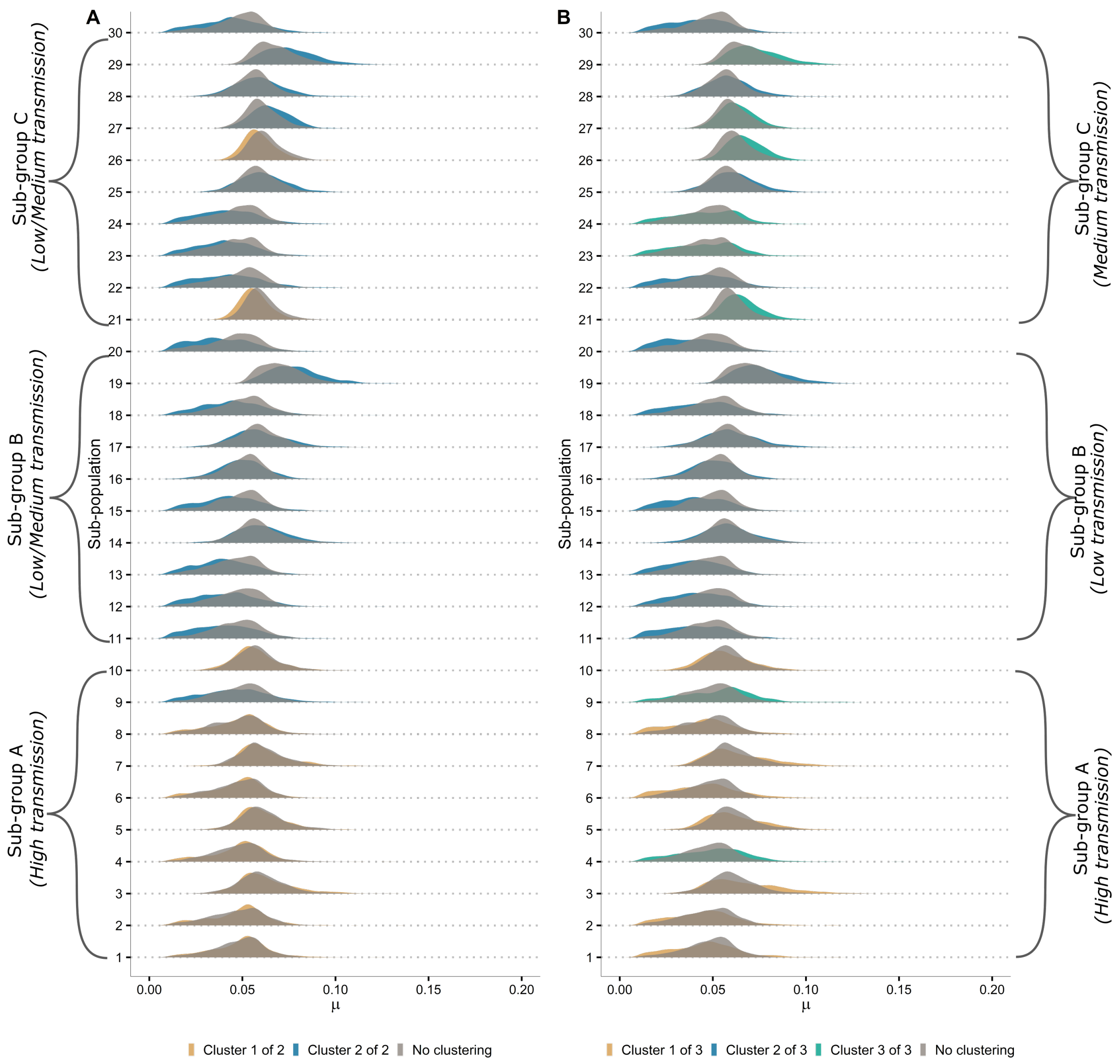


Figure S11: Marginal posterior distributions of $\mu_k, k = 1, 2, \ldots, 30$

### S4 Assessing the parameter estimates of sub-populations under different estimation regimes

The percentages of which the HPD interval calculated under different ROPEs are included as CSV files in GitHub (see below for the link).

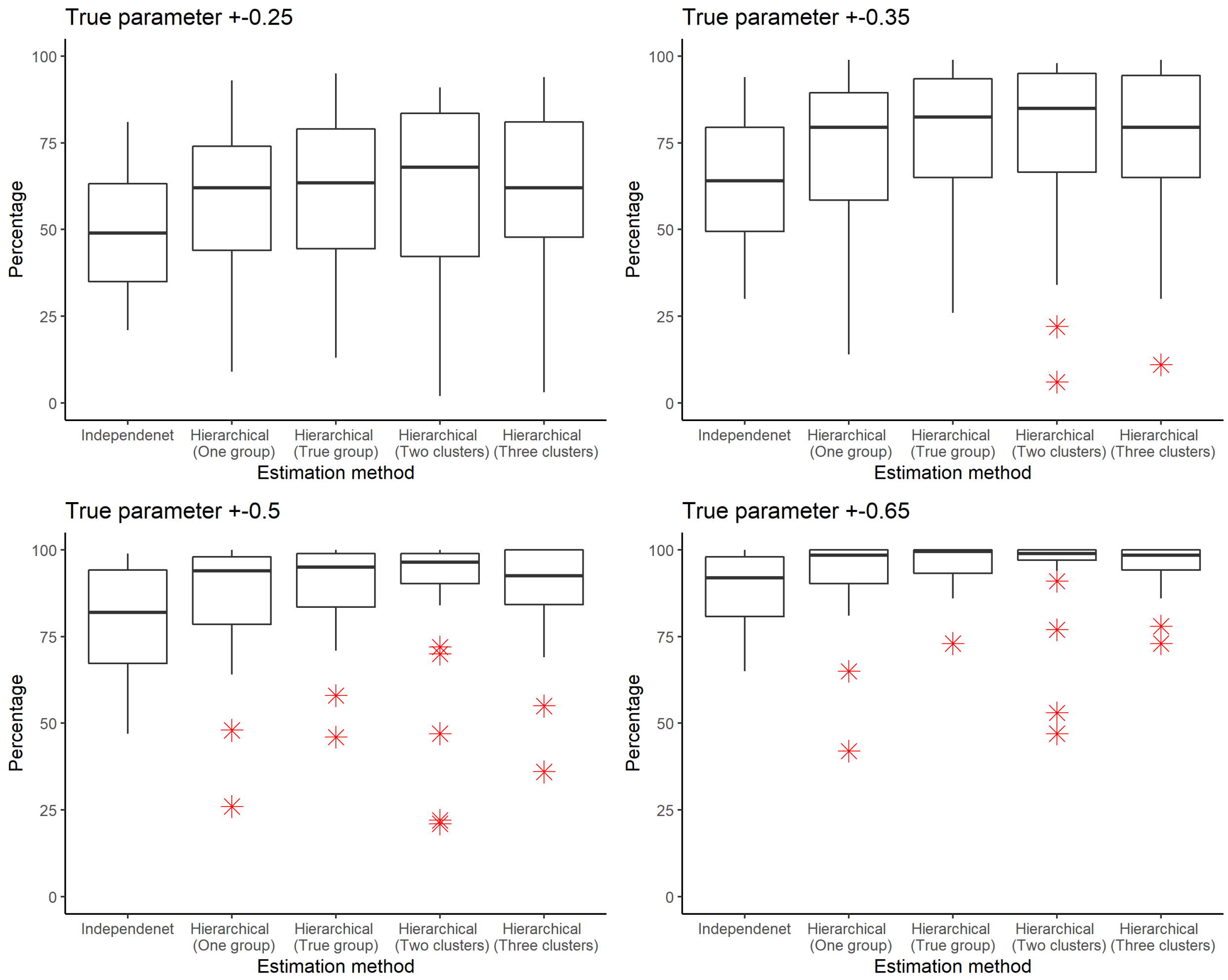


Figure S12: Box plots of the percentage of the 95% HPD intervals that intersect the ROPE intervals taken as observations across the 30 sub-populations under different estimation methods. Different $c$ values

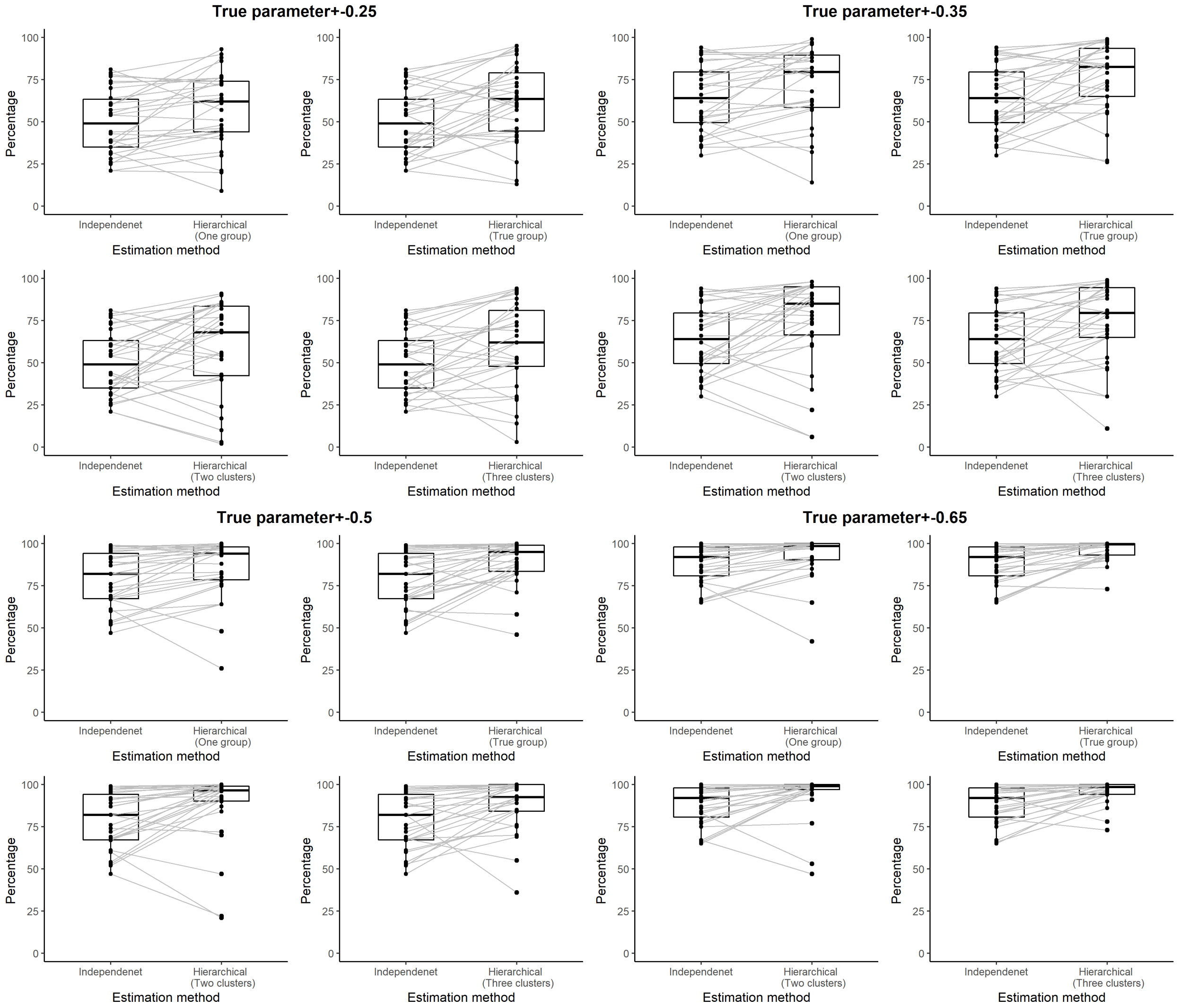


Figure S13: Box plots of the percentage of the 95% HPD intervals that intersect the ROPE intervals taken as observations across the 30 sub-populations under different estimation methods. Grey lines are corresponding sub-populations under each estimation method.

### S5 Criteria to identify an outbreak

The criteria we used to identify an outbreak is similar to that of Alahakoon et al. (2022a).

We generated a deterministic time plot for an *SIRS* model using the hyper means for each sub-group. We assumed these time plots as the general trajectories for outbreaks in each group. Accounting for the variability in each group and the stochastic effects, we then considered that the general peak prevalence of an outbreak for sub-populations in all the sub-groups will be greater than 50. We further assumed that for sub-populations in Sub-group A, once the first outbreaks die out, the trough will lie between the 5th and 20th days. For Sub-groups B and C, we assumed it will be between 10 and 25 days. When generating a sample path for a sub-population, if the trajectory did not have a peak greater than 50 or it did not enter the trough depending on the sub-population it belong to, we rejected that sample path and re-simulated until the criteria were satisfied.

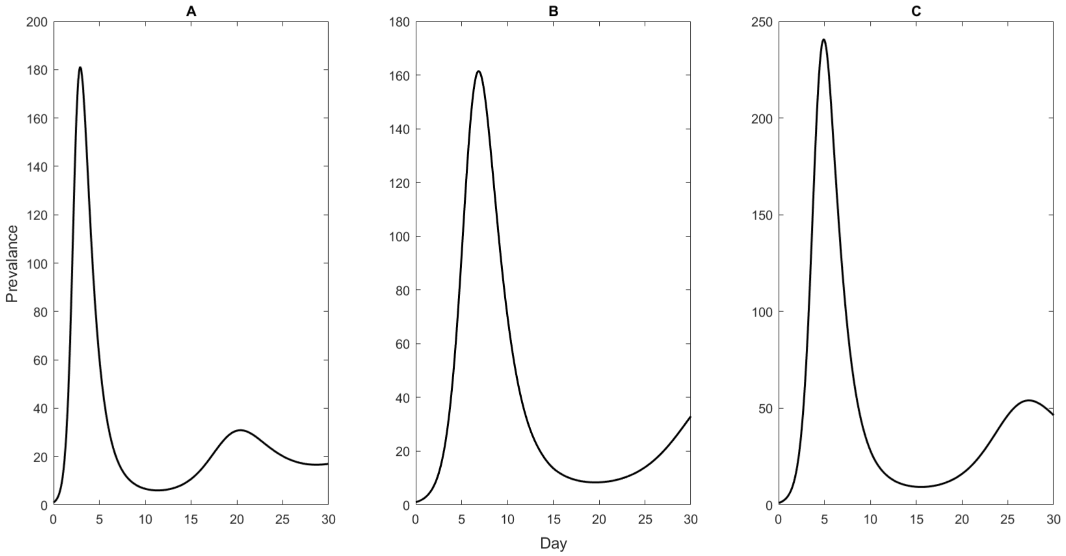


Figure S14: Deterministic $SIRS$ with parameters as hyper-means of Sub-groups A (panel A), B (panel B), and C (panel C).

#### S5.1 MATLAB Codes and related files

The codes and the related files can be found on GitHub at: `https://github.com/PunyaAlahakoon/Use_of_Clusetring_in_hierarchical_models.git`
